# The acceptability, adaptations, reach, barriers and enablers to the roles of endometriosis nurses and pelvic health & wellbeing coordinators in the Welsh Health Boards: a service evaluation by qualitative methods

**DOI:** 10.64898/2026.01.05.25341955

**Authors:** Eleanor Clarke, Natalie Rees, Denitza Williams, Alison Cooper, Libby Humphris, Adrian Edwards, Natalie Joseph-Williams

## Abstract

Endometriosis Nurse and Pelvic Health and Wellbeing Co-ordinator (PHWC) roles were introduced in each health board across Wales in early 2020. They were established to improve endometriosis and pelvic health care, as women can face severe problems and with major impacts across their quality of life, requiring expert care. How these staff roles have been incorporated into each health board has differed according to local need and as such, is not fully understood, nor are the benefits of the role as perceived by staff. The aim of this service evaluation is to explore the views and experiences of endometriosis nurses and PHWCs on those roles and services.

Mixed qualitative methods were applied (interviews and workshop) between January 2025 and June 2025 to explore the study objectives. Interview data were thematically analysed to identify key themes. Workshop data were reported. The Cardiff University School of Medicine Research Ethics Committee waived the requirement for ethical approval as the project was found to be a service evaluation.

The roles are important to health boards and post-holders with discussion of positive impact on patients across the health boards. Key factors in the success or limitation of these roles include:

**At the macro-level:** Availability of resources for staffing, training, administration, clinics and theatre time.

Service fragility: The services, particularly endometriosis, rely on usually one person per health board and so cease during leave or recruitment, with no succession planning.

**At the meso-level:** Colleague, peer and other healthcare professional support.

**At the micro-level:** Clarity of role description and establishing boundaries of the service. Support and guidance for new endometriosis nurses.

**Funding Statement:** The Health and Care Research Wales Evidence Centre is funded by Health and Care Research Wales on behalf of Welsh Government.

**Executive Summary:** *Background:* Endometriosis Nurse and Pelvic Health and Wellbeing Co-ordinator (PHWC) roles were introduced in each health board across Wales in early 2020. They were established to improve endometriosis and pelvic health care, as women can face severe problems and with major impacts across their quality of life, requiring expert care. How these staff roles have been incorporated into each health board has differed according to local need and as such, is not fully understood, nor are the benefits of the role as perceived by staff. The aim of this service evaluation is to explore the views and experiences of endometriosis nurses and PHWCs on those roles and services.

*Methods:* Mixed qualitative methods were applied (interviews and workshop) between January 2025 and June 2025 to explore the study objectives. Interview data were thematically analysed to identify key themes. Workshop data were reported.

*Key Findings:* The roles are important to health boards and post-holders with discussion of positive impact on patients across the health boards. Key factors in the success or limitation of these roles include:

1. At the macro-level:

- Availability of **resources** for staffing, training, administration, clinics and theatre time
- Service **fragility**: The services, particularly endometriosis, rely on usually one person per health board and so cease during leave or recruitment, with no succession planning
2. At the meso-level: Colleague, peer and other healthcare professional **support**
3. At the micro-level:

- **Clarity** of role description and establishing boundaries of the service
- Support and guidance for **new** endometriosis nurses

*Key Priorities:* Key priorities that could support the endometriosis nurse and pelvic health wellbeing coordinator roles are summarised in figure one below. These are summarised from table seven (section five). Figure one:
Summary of what could be done to support the endometriosis nurse and / or pelvic health wellbeing coordinator roles

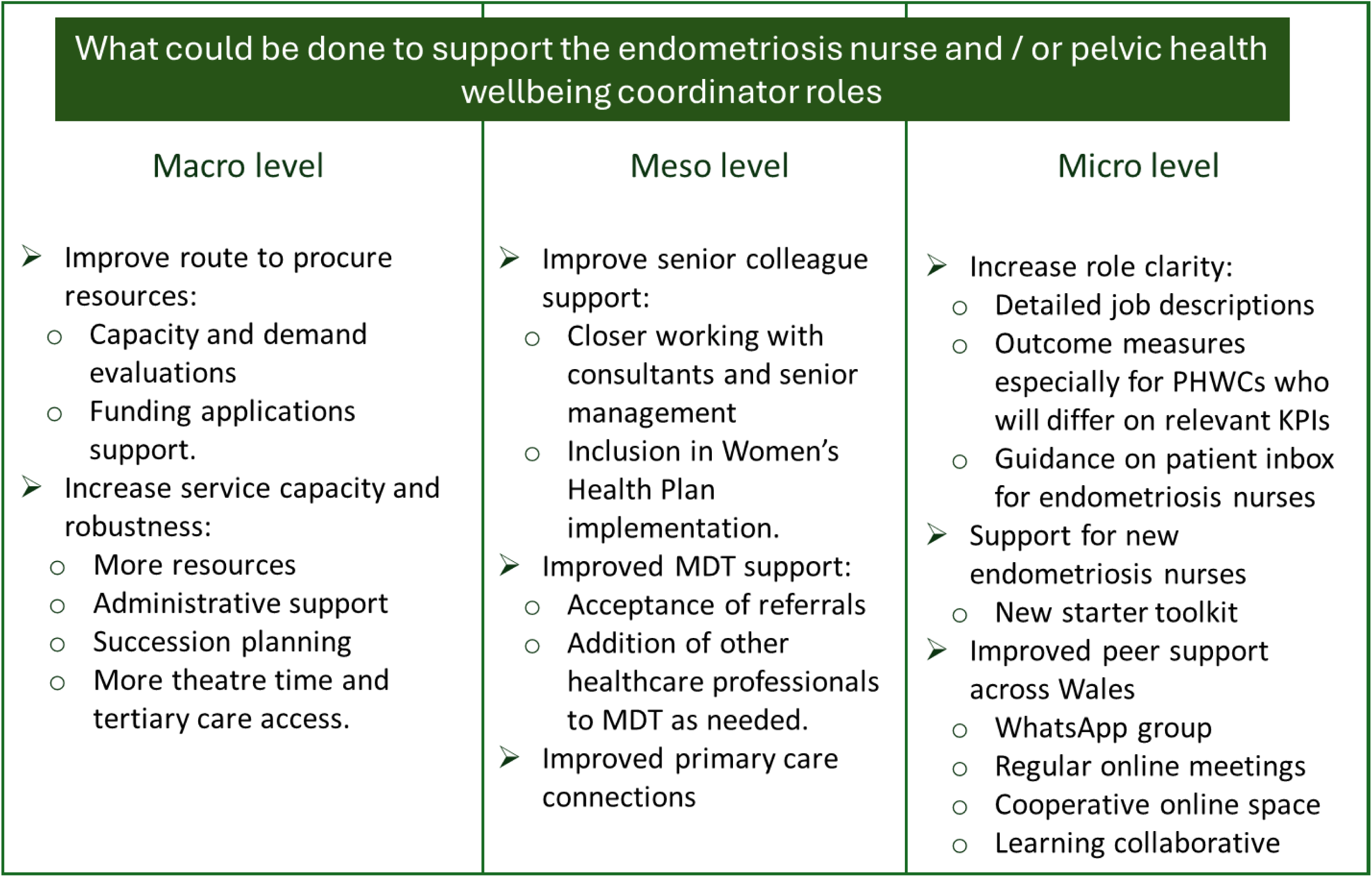

## 1. Background

The Women’s Health Implementation Group (WHIG) was established in May 2018 to provide leadership and strategic direction in relation to several key areas of women’s health policy where particular opportunities for improvement had been identified. Its remit was ministerially-driven and included ensuring an all-Wales approach was in place to help break down barriers, join up pathways between primary, secondary and tertiary care and manage women’s health in the community.

The initial priority of the WHIG was to oversee the implementation of recommendations from the vaginal mesh and tape review, however the Minister for Health and Social Services subsequently directed the group to also consider the recommendations arising from the endometriosis and faecal incontinence reviews. The WHIG has now been replaced by the National Strategic Clinical Network for Women’s Health.

Endometriosis Nurses and Pelvic Health and Wellbeing Co-ordinator (PHWC) roles were introduced in each health board across Wales around five years ago, and health boards receive funding from Welsh Government on a recurrent basis to support these roles. They were established to improve endometriosis and pelvic health care (including pelvic mesh complications), to improve access to expert care because many women can face severe pain and symptoms that impact their quality of life [1].

### 1.1. Endometriosis service

Endometriosis is a common gynaecological disorder, affecting up to one in ten women of reproductive age in the UK [2]. It is a condition where endometrium-like tissue is found outside the uterus, often in the pelvis (e.g. ovaries, peritoneum) that ranges in severity. Hormonal changes of menstruation then cause bleeding, chronic inflammation, pain, and scarring. The exact cause is unknown, and diagnosis can be a long, difficult path for patients, taking an average of ten years and 26 doctor visits from initial presentation to diagnosis [2]. Treatment includes symptom management, hormonal treatment [3] or surgery [4].

Through the Women’s Health Implementation Group (WHIG), specialist endometriosis nurses were appointed in each health board in Wales in Spring 2020 funded by Welsh Government [1]. A central job specification was developed, and each health board submitted a bid that requested support to meet local demand and capacity. The WHIG also set up the ‘[Endometriosis Cymru](https://endometriosis.cymru/)’ website to raise awareness and provide resources.

### 1.2 Pelvic Health Wellbeing service

There is an increasing focus on prevention and conservative therapies in the treatment of pelvic health problems. This may include pelvic pain, prolapse or incontinence. Aligned to this are complications arising from vaginal mesh surgery which require tailored and expert knowledge to support sufferers.

Pelvic health problems can include complications from vaginal mesh implants previously inserted during surgery for urinary incontinence, pelvic organ prolapse or birth trauma [5]. Concerns were formally heard in the ‘Review into Vaginal Mesh and Tape in Wales’. One recommendation from this review was that a ‘*care co-ordinator type role should be embedded within the Pelvic Health and Wellbeing Pathway for women with mesh associated pain as a first point of contact*’ for patients [5]. These co-ordinators would support the development of a Pelvic Health and Wellbeing Pathway from the community into tertiary services as necessary for other pelvic health issues such as continence care and endometriosis.

The WHIG introduced Pelvic Health Wellbeing Co-ordinator (PHWC) roles in each health board, funded by Welsh Government. Basic requirements of the roles included:

- Appropriate experience of working in the NHS.
- Support the Senior Programme Manager to implement a pelvic health and wellbeing care pathway in the health board from community up to, where necessary an MDT including continence care, physiotherapy, pain management and psychology services. For women and men.
- Undertake an assessment of any additional resources required for the pathway.
- Provide leadership and strategic direction to ensure the health board meets the full requirements of the Pelvic Health and Wellbeing Pathway.
- Sign post patients and facilitate ongoing care in the health board for pelvic health issues associated with vaginal mesh, endometriosis and faecal incontinence.
- Support the senior programme manager to provide expert advice to the WHIG, ensuring key health board stakeholders are engaged in the pathway design and delivery.
- Work with other PHWCs to form a Network to share best practice and identify opportunities for continual service improvement
- Work within the context of the health board governance arrangements.

Each health board then developed individual job descriptions and business cases based on local needs and submitted to Welsh Government. Staffing resource included in the bids varied from one to four members of staff; staff disciplines and hours requested also varied. These bids were signed-off and appointed during 2019.

### 1.3 Current understanding of women’s services in Wales

How these roles have been implemented, adapted, and incorporated into each health board service is not fully understood, nor are the staff perceived benefits of the roles. Further, no previous evaluations have examined key barriers to the roles being implemented in routine practice or summarised what is working well.

Evaluation of these roles can inform service improvement and implementation of future womens health services or roles across health boards.

### 1.4. Aims and Objectives

The aim of this service evaluation is to explore the views of endometriosis nurses and PHWC’s on those roles and services. We used qualitative interviews underpinned by process evaluation methodology and implementation frameworks to explore:

- What the roles look like in practice (and any variation from the intended roles)
- Acceptability of the roles (as defined by Sekhon et al [6] is the extent to which the participants consider the roles to be appropriate, based on their cognitive and emotional responses).
- Whether participants believe the relevant patients are being reached
- What has worked well and what supports the roles and service delivery
- Barriers and key challenges to delivering the roles and service delivery
- Recommendations for improvement going forward
  - Within the role
  - Across health boards
  - For other women’s health services

The process evaluation element was intended not only to capture what was happening in terms of delivery and perceived benefits but also to explore discrepancies between expected and observed delivery and outcomes, how context influenced these, and to provide insights for future improvement and sustained roll-out.

## 2. Methods

Mixed qualitative methods were applied (interviews and workshop) to explore the views and experiences of endometriosis nurses and PHWCs on their roles and service, including the barriers, facilitators and benefits.

Interviews focused on experiences and personal views about the endometriosis nurse and PHWC roles and services. During the workshop findings to date were shared and then participants suggestions for positive ways forward to support development of implications for practice for these roles.

### 2.1 Sample

The interview sample included endometriosis nurse and pelvic health and wellbeing coordinator roles (see Table 1).

**Table 1:**
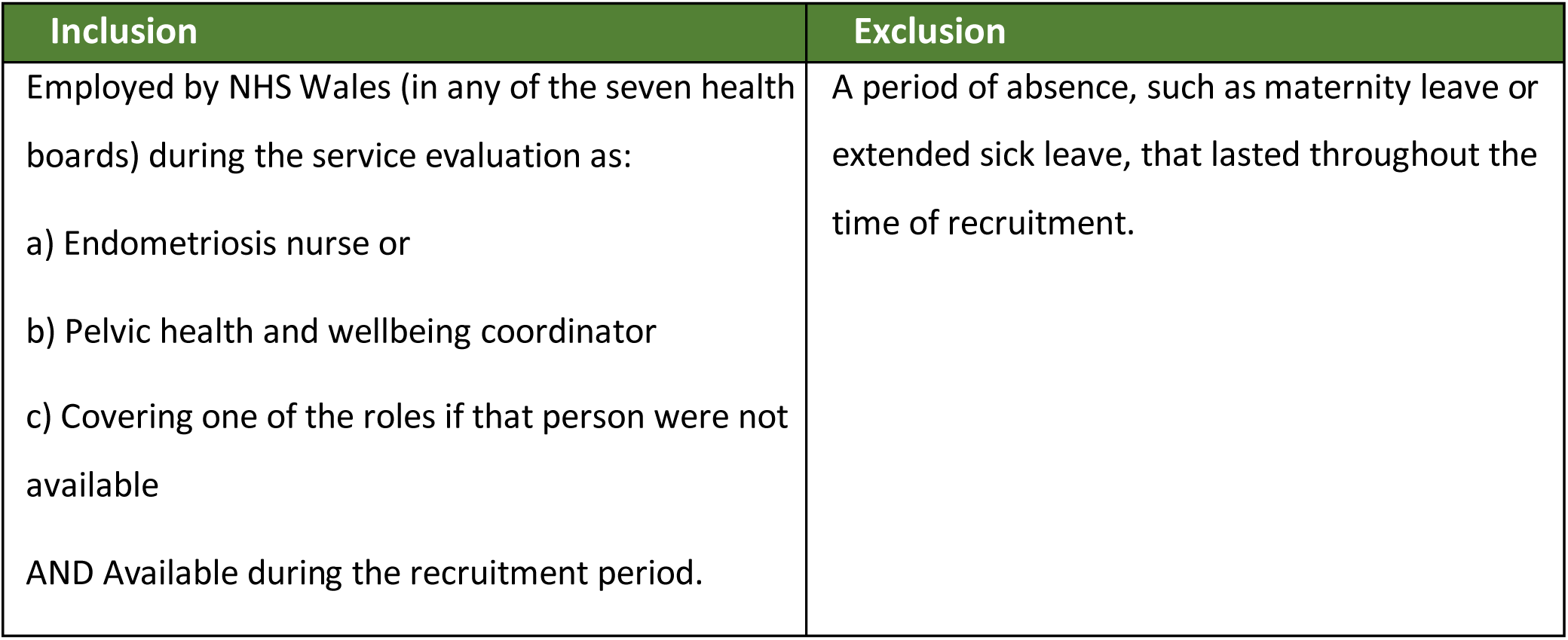
Interview eligibility criteria.

Those eligible for the workshop were all interview participants, study stakeholders, specialist interest women’s health researchers (limit of two slots).

### 2.2 Participant recruitment

#### 2.2.1: Interview recruitment

Targeted sampling was applied to invite all potential participants. The research team was provided with potential participants’ names and work email addresses. After receiving approval from each health board, potential participants were invited to participate. For those who agreed, online interviews were scheduled at a time convenient to them. A link to the electronic consent form (Appendix two) was provided in advance of the interview. A member of the research team (EC) was available to respond to any queries by email and at the beginning of the interview

#### 2.2.2 Workshop recruitment

Interview participants provided consent to be contacted for the optional workshop at the end of the interview. The workshop date was arranged in consultation with stakeholders and scheduled during lunch time to support attendance. All potential participants were contacted with a workshop invitation (appendix four) including the date, time, explanation of what would happen and why. A member of the research team (EC) was available to answer queries prior to the workshop.

### 2.3 Data Collection

#### 2.3.1 Interviews

Semi-structured interviews using an interview schedule (see section 3.5 below and appendix one) were conducted online via TEAMs between January 2025 and April 2025. Interviews were conducted by EC and NR and audio/video recorded. Please see box 1 for a summary of key topics.

##### Box 1

**Summary of key topics explored in the Interview schedule**

- **About the role:** description, preparation, support, how it fits in the system.
- **Working with others**: do you work alongside other practitioners, who, colleagues attitudes and expectations
- **Patient experience**: Are the right people being reached, patient impact, patient feedback, patient needs/desires, patient access, unintended consequences
- **Changes to the role**: evolution of the role and why, could anything be improved, barriers, facilitators.
**- Recommendations**: What are they? What has worked well? Significant challenges or changes.

The interview schedule was based on two theories. The Implementation Outcomes Framework [7] provides researchers with outcomes to assess the success of implementation (or execution) of an intervention (for this evaluaiton, these new women’s health roles). The MRC Process Evaluations Framework a guide for a systematic approach to conducting a process evaluation, which is examining how an intervention is implemented and delivered, for a complex intervention such as these roles. The interview schedule was then developed with the research team, stakeholders and Public Partner (LH). After the first interview the interview schedule was redesigned to flow more easily, refine topics and reduce the number of questions – topic schedules are designed to be iterative.

#### 2.3.2 Workshop

Following the initial results presentation, participants were separated into breakout rooms: one for each role (see Appendix three for workshop facilitator guide). The facilitators guide comprised four main topics (with prompts) as summarised in box 2 below.

##### Box 2

**Topics explored in the workshop**

- **Role clarity** (PHWC) or supporting **new starters** (endometriosis nurses)
- **Colleague** support
- **Boundaries**
- **Resources** (finance)

Workshop questions were developed based on themes identified as both barriers and facilitators to implementation. These were chosen for their potential to be influenced (excluding fixed contextual factors) and were shaped with input from both stakeholders and the research team. One question differed between rooms. Questions and prompts were asked by facilitators from the Evidence centre in the break out room (AC and NR in the endometriosis nurse room and DW and EC in the PHWC room). Discussions focused on identifying positive steps to support staff in their roles and improve services for patients. After the breakout sessions, all participants reconvened for a feedback session to share insights and collaboratively develop ideas for moving forward.

### 2.4 Data Analysis

#### 2.4.1 Interviews analysis

Transcripts were transcribed verbatim and imported into NVivo (version 14) qualitative analysis software. Endometriosis nurse and PHWC interviews were analysed together, with meaningful differences discussed in the results section.

Transcripts were thematically analysed [8] by EC and NR using deductive and inductive coding, with deductive codes from the MRC Process Evaluations Framework [9]. Inductive codes are those which develop during coding from the data itself. Codes and analysis were discussed at weekly project meetings with the study team (NJW, LH, NR, EC and AC) to support reflection, keep the analysis targeted and develop implications. Codes were grouped into overarching themes which were charted into a framework (including verbatim quotes).

#### 2.4.2 Workshop analysis

The workshop was recorded but not transcribed. Reflexive notes were made by all facilitators from the breakout session. One researcher (EC) listened to all the breakout sessions and the feedback session and compiled the discussions around the main themes of the interview data.

### 2.5 Researcher Reflexivity

Qualitative research acknowledges that researchers influence the research at every stage (from design to analysis). It is therefore important to reflect on the perspectives and experiences we bring to this project. EC is an experienced qualitative researcher with a psychology and medical background. She has led women’s health qualitative work and process evaluations previously. She is a white, non-disabled, cis-female with personal experience of using the Welsh NHS. NR is a GP with a special interest in women’s health, working in the Welsh NHS since 2003 and in general practice in Wales since 2010. She holds qualifications in women’s health including the DFSRH (2009), and is a trained contraceptive implant and coil fitter (since 2018). She is a white, non-disabled, cis-female and a novice researcher currently working as an Associate Academic Fellow at Cardiff University.

### 2.6 Ethics and governance

This project was classified as a service evaluation rather than research, and therefore did not require ethical approval. This classification was confirmed using the Health Research Authority Decision Tool (https://www.hra-decisiontools.org.uk/research/) (developed for the Medical Research Council) and verified by the Cardiff University Joint Research Office. The research team liaised with each health board’s research and development department individually to obtain permissions to conduct the evaluation.

An online Participant Information Sheet / Consent Form in Microsoft Forms (appendix two) was emailed to all participants prior to interview. It was made clear that they did not have to complete the consent form and that there would be time for discussion prior to the interview.

At the start of each interview, and before recording began, participants were reminded that they could withdraw at any time, they did not have to answer a question if they did not want to and that we could stop for a break should they want.

## 3. Results

### 3.1 Participants

#### 3.1.1 Interview participants

For this service evaluation, all 17 eligible participants were successfully recruited. Of these only one was recruited under inclusion criterion c, which allowed for the participation of a colleague covering one of the roles if the substantive postholder was unavailable. In addition, three endometriosis nurses were in role but not eligible to participate. Table two below shows demographics of interview participants based on interview data.

**Table 2.** Interview demographic data: From interviews, so is best as can be estimated from the data.

|  |  | Endometriosis Nurses | PHWC's |
| --- | --- | --- | --- |
| Total number |  | 10 | 10 |
| Number eligible |  | 7* | 10 |
| Total number interviewed |  | 7 | 10 |
| Number of health boards represented |  | 6 | 7 |
| Job role when PHWC | Nurse | N/A | 3 (1 uses the PHWC role time not clinically but on service improvement) |
|  | Physiotherapist | N/A | 1 |
|  | Governance / administrative** | N/A | 1 |
|  | Administrator | N/A | 2 |
|  | Project manager | N/A | 1 |
|  | Management | N/A | 2 |
| Primary clinical area of focus | Endometriosis | 7 | Within mixed |
|  | Mesh | N/A | 2 |
|  | Continence | N/A | 2 |
|  | Urogynaecology | N/A | 3 |
|  | Colorectal | N/A | 1 |
|  | Mixed of the above | N/A | 2 |
| Length of time working in the role (incomplete data) (months) | Range | 4 – 60 | 9 – 63 |
|  | Average | 27.7 | 47.2 |

\*Three were on long term maternity / sick leave, \*\*Not a clearly defined role within the interview

#### 3.1.2 Workshop participants

The workshop was attended by four stakeholders, four endometriosis nurses and six PHWCs (table three). As the workshop was in June, some were on leave and there were some who wanted to attend but had previous commitments.

**Table 3:** Workshop participants.

| Role | Number attended workshop |
| --- | --- |
| Stakeholders | 4 |
| Endometriosis nurses | 4 |
| PHWCs | 6 |

### 3.2 Interview Themes

Seven key themes and 21 sub-themes were identified through the interviews (see Table four).

**Table 4:** Interview themes and subthemes.

| Main themes | Sub themes |
| --- | --- |
| A: Role Description | Endometriosis nurses |
|  | Pelvic Health and Wellbeing Coordinator (PHWC) |
|  | Lack of Role Clarity |
|  | Differences across health boards |
|  | Getting started in the roles |
|  | Fragile service |
|  | At capacity and pressure to perform |
| B: Support from others | Advocates and senior staff support |
|  | Peers working together |
|  | Multi-disciplinary working |
| C: Reach and inclusivity of roles |  |
| D : Resources | Finance |
|  | Administration and clinic space |
|  | Theatre time |
|  | Staffing and training |
| E: Skills and Boundaries | Boundaries |
| F: Context | Tertiary care |
|  | Patient group |
|  | COVID |
| F: Perceived benefits of the roles |  |
| H: Service Development | Implemented or imminent changes |
|  | Ideas for future service improvements |
|  | Recommendations for the roles |

#### Theme A: Role Description

Endometriosis nurse and PHWC participants were passionate and knowledgeable about their work. They discussed the roles as being positive for patients (see theme G: Perceived benefits) and for many these roles have ‘*evolved’* (a PHWC) from the initial idea and continue to do so.

##### A.1 Endometriosis nurses

Endometriosis nurses had a shared understanding of the condition they treat, their expected profession and clinical background.

> *“the aim of the role is to support the care and management of patients with a diagnosis or suspected diagnosis of endometriosis in [the health board].” (an endometriosis nurse)*

An endometriosis nurse participant explained the purpose of the service:

> *“to add support for them [patients] and also the GPs to manage their care until they’re seen in gynaecology. Once they’ve been seen by us, they can still come back […], if they need any help or support, they can still come to this clinic if they’re having symptom problems or just need a little bit of advice”.*

Some endometriosis nurse participants described what might happen at an initial appointment, which is usually face to face (see box 2). However, this is not exhaustive and would depend on the patient. They then follow-up with patients, usually virtually, in shorter 20-30 minute appointments (see figure two for an example of how an endometriosis nurse service can flow). The regularity of these clinics differs across health boards.

**Figure 2:**
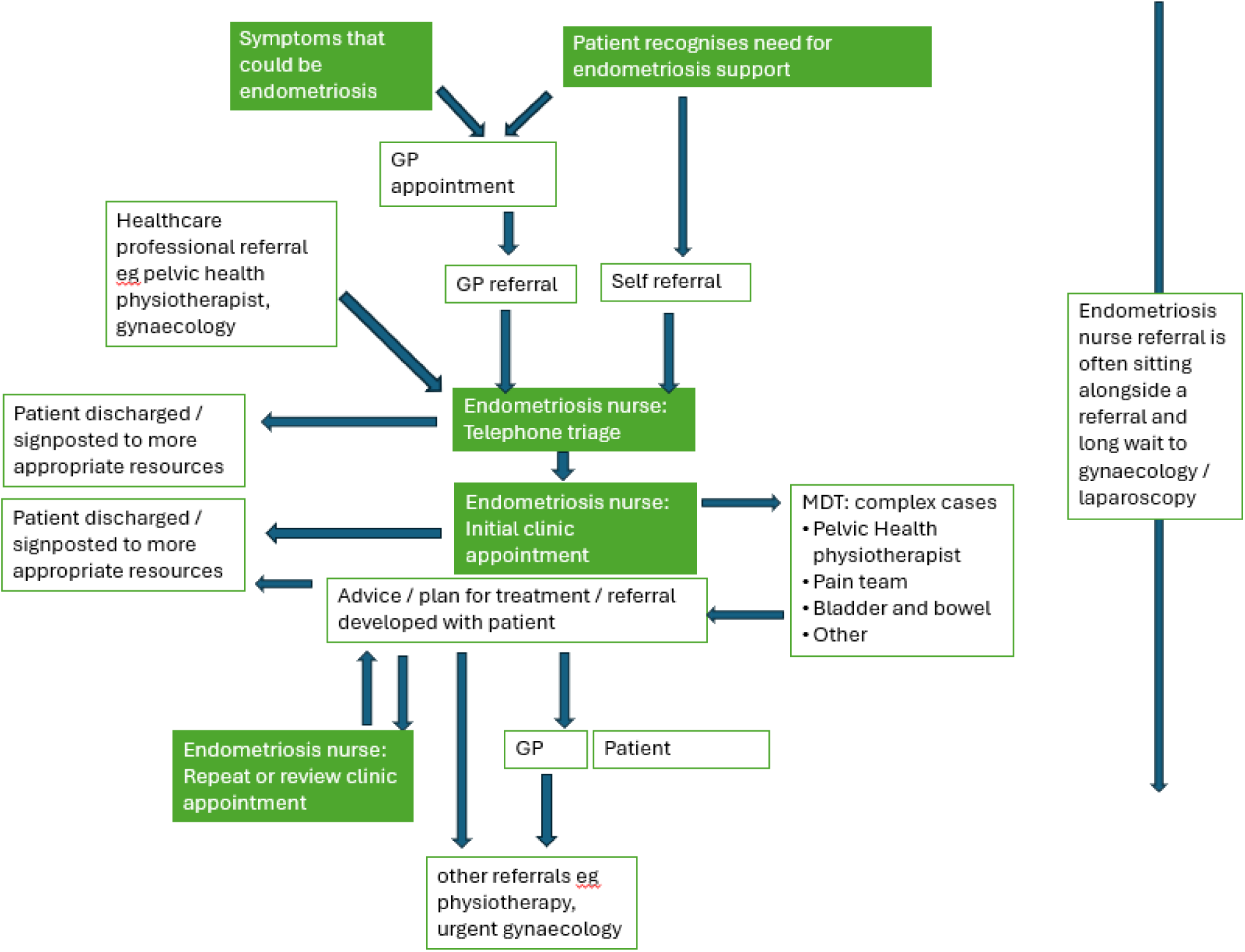
An example of a patient pathway through the endometriosis nurse service

###### Box 2

**What can happen at an initial endometriosis nurse appointment**

- Pre-appointment pelvic health pain questionnaire
- Medical and surgical history from the patient (despite often notes being provided). Extra focus on:

- Rich gynaecological history (e.g. menarche).
- Sexual function will be discussed
- Bowel symptoms
- Back pain
- Urinary symptoms
- Fertility wishes
- General energy
- Symptom discussion

- Body mapping (of pain)
- Pain scoring (to track)
- Impact of symptoms on daily life
- Treatment discussion: what has been tried so far
- Discussion of endometriosis diagnosis and / or prognosis

- Explanation of surgery and its likely benefit (or not)
- Sometimes there are tests such as:

- Ultrasound
- Smears
- Swabs
- Formulate a plan with the patient

- Discuss potential referrals / requests for tests
- Discuss hormone therapies - new / altered: may be initiated with the consultant. May be just information.
- Explain support, e.g. Endometriosis Cymru [10] and their symptom tracker [11]; Endometriosis UK [12]: links to be sent after appointment.
- Any need for a letter to an employer
- Clarification of what information will be sent after

Afterwards a clinical note is sent to the GP and patient with any information requested. Referrals, tests requests, letters, etc actioned.

Participants talked about listening to patient journeys and counselling them through the reality of prognosis and treatment options.

> *“a big part of my role is listening and being able to create a space where a patient can be heard, because they’ve been gaslit and think that a lot of them don’t trust health care professionals, because of their experiences.” (an endometriosis nurse)*

There were reflections that clinic appointments could be difficult for patients who come with different expectations.

> *“I will give them all the tools and all the information […] It’s up to them if they choose to do it or not. I can’t force it upon them, but I also give steps, so I’ll give them maybe three or four steps ahead, so that if this doesn’t work, they, they can try this.[…] it also empowers the patient with knowledge that they could go to their GP and say well, this is what they’ve said.” (an endometriosis nurse)*

Endometriosis nurses work beyond their own endometriosis nurse-led clinics. Other tasks discussed included: Supporting consultant-led clinics; attending general gynaecological clinics; pre or post op counselling; supporting patients in other appointments; perform post-operative follow-up; lead endometriosis MDT meetings; medication injection clinics; fertility clinics. Some endometriosis nurses also do their own administration which is time consuming.

> *“I do a lot of administration, which probably takes up about 40, 45% of my time, which will take me away from being patient focused quite a bit.” (an endometriosis nurse)*

Endometriosis nurses give patients a phone number or email to contact them at any time with their concerns or queries. This was reported as an important task but created “*an awful lot of work on a daily basis because it fills up”*.

##### A.2 Pelvic Health and Wellbeing Coordinators (PHWC’s)

> *“I think the main goals of my role is to try and improve, focusing on women’s pelvic health at present, how we can improve waiting times, treatments of patients and outcomes, how we can better reach these patients in a better timely manner.” (a PHWC)*

It became apparent during interviews that there was a real variety within the PHWC roles. This can be seen from the demographics in table two (section 3.1) above. Clinical areas varied from a focus on mesh to a focus on continence or more general urogynecology issues, with some services including male patients.

Profession of PHWCs varied from non-clinical, such as administrative and governance to clinical staff such as physiotherapists and nurses with some not patient facing.

**Figure 3:**
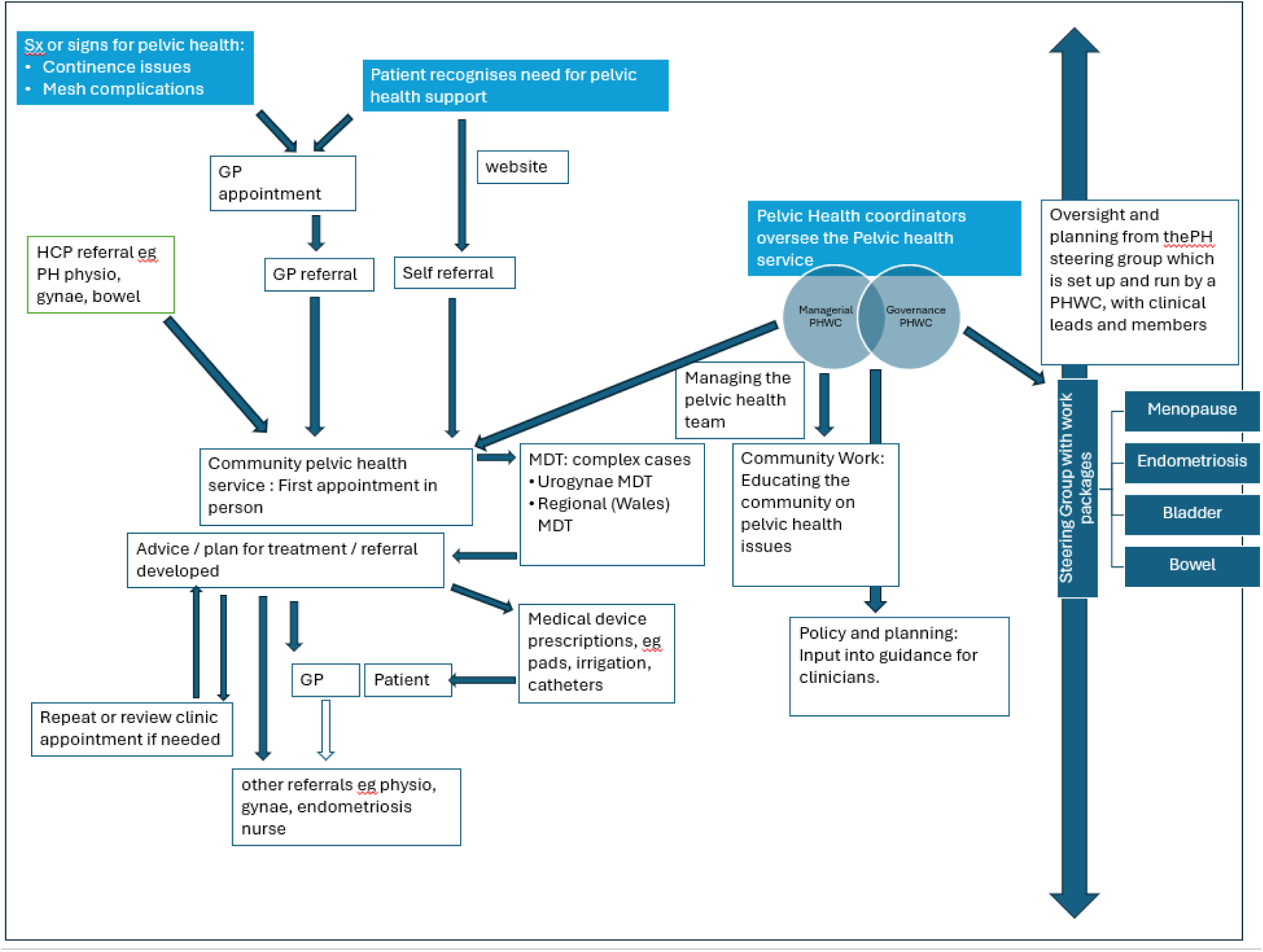
An example of a pelvic health and wellbeing service with two pelvic health and wellbeing coordinators *These services are incredibly varied. This example shows one service with two PHWCs who worked in different aspects of the role.*

Clinical PHWCs discussed in-depth history taking, examination of pelvic pain and standard data scoring to support surgery decisions such as the “*central sensitisation inventory*” (a PHWC) for mesh patients. Some participants described specialist therapies and clinics, such as one who offers a broad range of procedures and tests within urogynaecology.

Improving waiting times within pelvic care was seen as an important outcome of the PHWC role.

> *“we’ve been, sort of, steered more to look at urogynae side and waiting lists” (a PHWC)*

Some services were set up through identifying a need within the health board around pelvic health, perhaps giving long or short term aims. At least one participant felt that the PHWC role was to advocate for women patients within the health board.

> *“Everything around the mesh issues that I read […] part of the role, I thought,[…] was to actually coordinate as a health board with higher management and the stakeholders to improve care so things like that didn’t happen again.” (a PHWC)*

##### A.3 Lack of role clarity

Participants reported that the way in which the role is described and understood impacts their work. Lack of role clarity, particularly around the PHWC role, could cause confusion especially as the roles were being set up or if there was not hand over between staff.

> *“I think at that point, none of us really knew what, what we were starting with, where we were starting, or what was needed.” (a PHWC)*

PHWC roles differ widely in terms of the clinical area they focus on and the profession of the post holder (table two, section 3.1), where staff may not be patient facing, nor have a clinical background:

> *“the conversations we’ve had at the pelvic health network that there can be physios, there’s admin people, there’s nurses. […] that’s the, the tricky bit with this role I think, is that no one role is the same and so if you say it, it can mean a number of things to a number of different people in terms of what we’re actually doing in the service really.” (a PHWC)*

Participants discussed confusion not only in the definition of the PHWC role but within the title itself.

> *“the thing that I find can be confusing for people is the term ’pelvic health’ because, […] pelvic health covers such a wide area, doesn’t it. I mean, even just in women’s alone, you know, you can look at pelvic health in terms of maternity, postnatal, antenatal, you can look at it in gynae, and it can be endometriosis, it can be pelvic floor dysfunction, it can be menopause, menorrhagia.[…] So, I think that’s where the title has been misleading, I think. Or could be confused.” (a PHWC)*

Being in the same role title but working with different patient groups can be difficult for staff to manage.

> *“it’s a very confusing role for me as to what we’re supposed to be doing as Pelvic Health Coordinators. […]. So where we thought everyone was going to be setting up a service just to see mesh patients, it didn’t sort of come across as that when we were in these meetings.” (a PHWC)*

Another participant explained that amongst all the pelvic health issues that “*mesh […] that was a little bit forgotten from my perspective*” (a PHWC). In one health board at least, it was discussed that the PHWC role would have a different clinical focus,

> *“I think the feeling very much was that those [mesh] patients were already known to us and they’ve, they’d either been supported or they’d been then signposted to tertiary centres for resolution of their, you know, problems […], I think the feeling was that there was already a system in place. And I, if I’m honest, I don’t know that they were very clear” (a PHWC).*

Pelvic health is not a gender specific term, in some health boards the PHWC service treats men, in others they do not. This caused confusion or a lack of engagement from some staff or teams.

> *“Because the money came in to women’s specifically […] there wasn’t the, kind of, engagement, there wasn’t the same buy in” (a PHWC).*

Some participants, particulalry PHWCs, discussed that role clarity could be improved by introducing KPIs (Key Performance Indicators) or alternative outcome measurement. Other participants felt that would not be feasible due to the variety in the roles and because they were not set from the start.

> *“we haven’t had, as far as I am aware, any real KPIs or any clear steer on definitely what our outcomes would be. […] So, I think that’s what made it quite difficult really” (a PHWC)*

##### A.4 Differences across health boards and roles

Some key service differences found during interviews are shown in table five below. It is important to remember that this was qualitative data and not everyone was asked the same questions.

**Table 5:** some key themes of difference between services.

| Key emerged theme of difference |  | Endometriosis services* | PHWCs |
| --- | --- | --- | --- |
| Conduct own administration within clinical role |  | 3 out of 6 | 2 out of 5 |
| Referral into services | Patients are able to self refer | 2 out of 6, with a 3<sup>rd</sup> on hold. | At least two services, not all were discussed or relevant. |
|  | Need for a health board consultant already involved in the patients care | 2 of the 6 services discussed a need for patients to be under a health board consultant. | Not discussed. |
| Admission criteria: only patients with a confirmed diagnosis of endometriosis (not suspected) can be seen |  | 1 service discussed this | N/A |
| Onward referrals to physiotherapy declined due to lack of funding for the patient group |  | 3 services found referrals rarely or never accepted | Not discussed. |

*\*One endometriosis service was not included due to long term leave and so there are a maximum of six endometriosis services discussed.*

Differences between health boards that were relevant to the job roles were discussed by participants (see box 3).

> *“this is an extremely rural health board, huge ageing population that’s increasing […]. Huge challenges in terms or kind of locations.” (a PHWC)*

Some participants perceived that the roles had aimed for the same service across Wales.

> *“The initial plan was that everybody would develop the same services. I don’t think that’s happened.” (an endometriosis nurse)*

Participants are aware of these differences and discussed the difficulties this can have for them as well as the impact on patients.

> *“We have meetings with all the other Welsh endometriosis nurses, I know other people are not, have not been in a similar position and it’s really sad. Really, really sad for them and the patients, because it’s not fair that, you know, somebody can live over the, in another health board, and not get access to the same service that they could get here, you know. So, yeah, don’t know how we solve that.” (an endometriosis nurse).*

Hours for the roles differ between health boards, particularly for PHWCs, as these were determined by local health board needs and then bid for rather than specified. This means there is variation in what can be achieved.

> *“I’m only doing two days a week anyway, so what, what somebody can do on two days is not what somebody on five days can do” (a PHWC).*

###### Box 3

**Relevant health board differences (not service specific)**

- *Access to secondary/tertiary care*
- *Support for the service*
- *Ability to make change*
- *Population e.g. age, deprivation*
- *Urban vs rural geography*
- *Sparse vs concentrated geography*
- *Organisational change*
- *Transport links (for patients)*
- *Division of health board administratively*.
- *Access to resources*
- *Different computer systems / software*

##### A.5 Getting started in the roles

Some PHWC’s were slotted into existing services which utilised their previous skills and were able to start quickly. But for other PHWCs and the endometriosis nurses, there was a need to set up a new service.

> *“there was nobody to copy off or fall into so I just had to, sort of, develop it myself” (an endometriois nurse).*

There were sometimes local issues which delayed setting up the services, with one health board only being able to set up endometriosis clinics in 2024 which was “*out of our control as nurses. It was a bit higher up”* (an endometriois nurse).

For new starter endometriosis nurses there was a period between employment and starting independent clinic work, regardless of how long the service had been established. Participants explained that this was due to the uniqueness of the role and pressure for high knowledge and skills needed to run clinics independently. The period between starting the role and being confident to run clinics was up to six months.

After the initial intake of endometriosis nurses (who were given training), there is not a routine induction for staff in these roles. Participants discussed a variety of ways they had (or were in the midst of) prepared for the endometriosis nurse role, especially for independent clinics. Participants reported shadowing clinical staff over TEAMs and in-person, attending operating theatre “*seeing the disease and, you know, what it does.”* (an endometriois nurse).

> *“it really set me up, you know, observing them in clinic, and how they did their paperwork, the questions they asked, that allowed me, you know, to learn a lot and to then sort of mirror how I would do my clinics” (an endometriois nurse)*

Some PHWC’s discussed similar preparation activities to the endometriosis nurses, though one PHWC commented that, “*a lot of it is self-taught, I will, would say*”.

##### A.6 Fragile service

During recruitment, there were two health boards where the endometriosis nurse led service was not running and at least one other where it was reduced, due to staff on long term leave. We were unable to interview anyone from one of these services. Participants discussed the fragility of their services (for PHWCs this concern of service fragility depends upon their role) where there is often one person running the specialist service for the entire health board or a large geographic section.

> *“It’s such a fragile service and we need more endometriosis nurses to be able to cover each other, you know. It’s having just one person is, is not enough […] It’s not a robust service at all.” (an endometriois nurse)*

> *“We’re just so fragile and, you know, small that until we can develop we are really restricted I think. […] basically, if, like, the nurse’s off sick or whatever there is no service.” (a PHWC)*

There is concern from participants at the lack of staff able to take over and cover the role as well as awareness of impact on patients.

> *“I’m very proud of the work that the endometriosis nurses are doing. It’s really valued and very needed, which goes to show that when the nurse is taken out of the equation, what a void that’s now created. […] They’re [patients are] desperate. They’re desperate and I completely understand. They need someone.” (an endometriois nurse)*

There was discussion that part-time job shares could attempt to cover for each other but that can be too difficult if the role is split geographically. Providing cover for these specialist roles is not straightforward, even if it is planned leave (such as maternity leave).

> *“we haven’t got cover for her, because it takes about six months for me to train someone. By the time we train a nurse to, to fill that role and all the checks, et cetera have gone through she will be back.” (an endometriois nurse)*

##### A.7 At capacity and pressure to perform

Participants are aware they are running a much-needed service for a patient group who have often been let down by other health professionals. Despite most wanting to reach out to more potential patients there is concern that there is not capacity in the service for increased demand.

> *“how much do we advertise the service right now to open the gates and just let them flood in. Or do we just try and try to, try to titrate it in a little bit, bit by bit, just so we don’t get too completely and utterly overwhelmed?” (an endometriois nurse)*

Participants don’t always feel equipped to manage all that is being asked of them. The high demand is felt keenly by participants, who need more resources to cope (see resources theme) as they see firsthand the effects on patients.

> *“We can’t say to people you’re waiting three to four years for surgery, but then […] not offer them the support that they need […] I’m only one person, and I’m not qualified in all of that. I can do as best as I can, but I think we have a duty to these ladies that we need to support them, and at the moment, I do feel we’re not giving them that service.” (an endometriosis nurse)*

#### Theme B: Support from others

Participants reported that support from others (including the MDT, line managers, peers and other health board staff) was an important factor in delivering the role. Levels of support vary across Wales.

##### B.1 Advocates and senior staff support

Some participants reported having a member of staff as an advocate, who may be more senior. This was described by one participant as a ‘*driving force’* and these advocates or mentors could offer encouragement, advice, contacts, funding or support with seniors. They contributed to an environment where the service was encouraged to grow and develop. Whereas other participants felt more isolated and shouldered the work alone.

> *“got [name 1], she’s a force to be reckoned with. She really is. […] she’s the driving force behind it. […] she will kind of give me something and say, […] ‘run with it’.” (a PHWC)*

Participants discussed relationships with senior staff as potential barriers or facilitators to their work. Consultants were discussed as key stakeholders:

> *“the ones who aren’t doing that great I can categorically tell you is because they don’t have the support of their consultant.” (an endometriosis nurse)*

Line managers coud be from different professional backgrounds which participants reported could lead to feeling misunderstood or unsupported. One participant reflected on having a line manager of a similar background:

> *“she’s the first line manager I’ve had […] that’s got experience in women’s health. […] she’s very focused on the endometriosis service, and it’s one of her main focuses as part of her new role. […] she’s very invested in making the service bigger and better” (endometriosis nurse).*

Perceived support or interest from senior managers was important to executing role tasks, particularly changing services.

> *“when we wanted to create this [new service], our directorate management team were very supportive” (a PHWC).*

Senior management interest is seen as an indicator of the prestige of the service within the health board. Participants, particulalry PHWC’s, reflected that senior managers may be more engaged if KPIs (Key Performance Indicators) or other outcome measures were linked to the roles.

> *“my higher management don’t have a clue what we’re actually doing in the service. […] if you’re not measured it does, kind of, start to go, ‘Oh, it doesn’t matter, does it?’ Whereas if you are measured, you have to keep delivering.” (a PHWC).*

##### B.2 Peers working together

Participants shared resources between peers to support each other and reduce health board differences (theme A.5). However, this approach did not seem to be formalised.

Participants discussed meeting up regularly for each role. The PHWC’s explained that this no longer takes place but could be beneficial to restart to support peer working and sharing of good practice.

> *“I’m quite passionate that I think from an All Wales, we need to be working together. Even if it’s just as coordinators. We used to have meetings, just the coordinators. We don’t even do that. [Interviewer: Since COVID or?] Since COVID, yeah. And they, you know, it was just to share ideas; share the governance that you’re using. […] because [this health board] is unique and, you know, there are different things, but the basics should be similar. You should all adapt and be pretty much using the same thing.” (a PHWC)*

Conversely, some endometriosis nurses discussed still meeting though it is unclear how well utilised this meeting is: “*Every fortnight, I think it is, but I haven’t been to one yet*” and “*I can’t comment on whatever they, what they’re doing in other health boards. I don’t know.”* mentioned another endometriois nurse.

##### B.3 Multi-disciplinary working

The NHS works on a model of multi-disciplinary care and so participants working clinically refer (or need to refer) patients to other healthcare professionals so that patients can receive holistic and appropriate care. Those whose referrals are accepted report being able to provide better care for patients. However, some participants report that referrals to some healthcare professionals are rejected due to funding and contract decisions. As shown in table five (section A.4), three partcipants discussed that referrals to physiotherapy are often declined. This is a barrier to care for patients, and can result in the participant having “*a bit of a battle*” to access those services.

> *“Physio management level, there’s no funding.[…] and they haven’t got the capacity in their service, with their current staffing to do it. It’s not that they’re being difficult. They want funding” (an endmetriosis nurse).*

An outcome of multidisciplinary working can be mixed messaging for patients. For example one participant explained that consultants could make surgery seem more positive than other members of the team would describe it. For patients this could be confusing and make other conservative treatments seem less worthwhile.

Endometriosis nurses discussed wanting to support GPs manage patients in the community through (1) promoting their service and (2) training GPs on endometriosis diagnosis and treatment. Participants reported struggling to contact GPs when they had tried to reach out.

> *“I think access to primary care is one of my biggest barriers.” (an endometriosis nurse)*

There was discussion of GPs supporting care plans, but then an example of a GP changing a care plan:

> *“a patient being given the hormonal contraception and told to take it for three months, try not to have a period. […] She goes to the GP, […] who said, but you’re a woman, you are supposed to bleed. So this poor girl then spent the next few months still going through the same thing, going completely against what she’d been advised. To me, that’s quite worrying, for one, because I think actually, you’re going against what has been advised.” (an endometriosis nurse)*

#### Theme C: Reach and inclusivity of the roles

Participants reported varied success in how much they felt able to reach the right patients and different communities. Some explained that to reach more patients there would need to be more staff in the role

> *“we see people, different, different localities, you know, different needs […] Which is nice, because everyone is accessing our [clinical] service.” (a PHWC).*

> *“There probably could be more, but there needs to be more of me.” (an endometrisis nurse)*

Some felt that there were no missing patient groups, whereas others reported less reached patient groups. Examples included younger patients, those from ethnic minority groups, those who can not speak english, people without a fixed address, the traveller community and men.

Some participants discussed that **self-referral** for women’s health services could or did improve access for people from different communities.

> *“especially women’s health, […] you don’t want everybody to know, do you? And in small communities like this, unfortunately, sometimes you can’t get away from that. So I really do think the self-referral element is important” (an endometriosis nurse)*

Of the thirteen services discussed in this service evaluation, four (table 4) were reported as having a self-referral route in interview data.

Service promotion varied, from working with colleagues to further outreach such as a PHWC who explained different outreach initiatives such as coffee mornings.

Participants reflected that there was some uncertainty about reach partly because there is a lack of relevant data in some services, with one service working on this issue at the time of data collection.

> *“We’ve got a study going on at the moment where we’re asking patients just to fill out a quick questionnaire, just so we can demonstrate that most of our patients coming through are white and British so that we can go and say, can you give us some funding, or something that we can start to integrate?” (a PHWC).*

However, some participants reflected there is not the capacity to collect extra data.

> *“the nurses approached us last week and they want us to fill in this database thing for patients coming through the door […] And this database would really benefit us just to show how many patients we’re actually dealing with. But […] we don’t really have time to capture that in the first place.” (a PHWC).*

#### Theme D: Resources

Availability of resources varied, especially for clinic space, MDT and administration support. Participants discussed the areas of women’s health and endometriosis as generally under resourced.

##### D.1 Finance

Participants generally discussed funding with a weariness: “*It is difficult because when we suggest things it’s always comes back to funding and money and that is a major, sort of, set back*.” (a PHWC). Participants are frustrated at what the lack of funding into women’s health indicates, as one endometriois nurse explained: “*because it’s [endometriosis] underfunded, it’s under prioritised*”. There was discussion that women’s health and endometriosis specifically struggle to get funding partly due to a lack of evidence and recognition in the health service:

> *“Because, you know, it’s one, this disease is everywhere, but it’s not recognised. We need to raise it and let everyone see it costs more than diabetes to society. […] there’s no NHS cost for endometriosis on […] that I can find.” (an endometriosis nurse)*

Participants seemed generally unsure how to increase funding and resources, with some considering if allowing service overwhelm would ultimately lead to service expansion:

> *“The only way we will be able to develop the service is by literally having a waiting list and then actually realising, ’Okay, you’ve got demand. We will find some money and help you.’ That, that’s the only way we’re gonna do it. We’re gonna have to be swamped to expand.” (a PHWC)*

Another participant reported they did not have the skillset of knowledge to produce a business plan independently:

> *“Truthfully, I wouldn’t even know where to begin with a business plan. So this is why I am working with [colleague]. But again, I fall short. I don’t even know where, who you would contact” (a PHWC).*

It was reported that England felt better resourced than Wales

> *“England’s got 64 endometriosis centres. We’ve got, what… Well, one. [health board] tryna be one, we’re tryna be one, but we’re just getting blocks because of money” (an endometriois nurse) [researcher note: currently Singleton hospital in Swansea and UHW in Cardiff have BSGE accredited endometriosis centres with a provisional centre at the Grange in Newport and a private centre at the Spire in Cardiff: bsge.org.uk 15th July 2025]*

##### D.2 Administration and clinic space

Space for clinics and provision of administrative support for clinical staff varies between health boards. Clinical staff who do their own administrative work then have less time with patients.

> *“we’re very lucky here in [health board] to have the service that we’ve got. And I know that a lot of the other pelvic health coordinators who are clinical, they struggle with the admin side of things” (a PHWC).*

Clinical staff are also not trained in administrative tasks and so take longer than a specialist.

> *“I don’t have any admin support. So I type all my own letters, book all my own clinics, and obviously do all my own leaflets. I do a lot of administration, which probably takes up about 40, 45% of my time, which will take me away from being patient focused quite a bit.” (an endometriosis nurse)*

Clinic space is not a barrier for some participants and for others is prohibitive. For many, virtual clinics have alleviated some of this pressure.

> *“We have to fight for it, continually fight for it because of other services.” (a PHWC)*

##### D.3 Theatre Time

Restricted operating theatre time and resulting long waiting lists for surgery are a major issue for all endometriosis nurses and some PHWCs by limiting the care they can provide within the service.

> *“You need more theatre time. We have a huge waiting list.” (an endometriosis nurse).*

Participants discuss being a support for patients while they endure a long wait for surgery which can be effected by local decisions.

> *“[A surgeon] lost when she went on maternity leave, her theatre list was given to somebody else, and they refused to give it back […] You need more theatre time. We have a huge waiting list” (an endometriosis nurse).*

A lack of theatre access is compounded by cancellations:

> ***“****if lists get cancelled, and these ladies who’ve been waiting three years could then potentially wait another few months. So I’m, I’m constantly firefighting patients, trying to apologise or I’m sorry about the waiting list, whereas if we were operating on people in a more timely fashion we could be getting through a lot more”. (an endometriosis nurse)*

One PHWC explained that the theatre time for their service increased through business cases compiled by the consultant (advocate theme B.1).

> *“when I started the role, we struggled to get theatre time and theatre capacity for these patients. As a driving force of [name 1 - consultant colorectal surgeon], she’s managed to get it funded by the health board, and we had to do business cases, and we had to present it to the clinical board and things.” (a PHWC)*

##### D.4 Staffing and training

Staffing is an issue for participants, with discussion of a need for more staff in the same role (i.e. more endometriosis nurses and PHWC’s). Participants report being unable to meet the needs of patients (related to theme A.7). There is also a need for more staff to cover leave (see A.6 Fragile Service).

> *“with one in 10 women diagnosed or queried endometriosis. I just, I was just scratching the surface. There are so many people to see and I did feel I wasn’t in the best quality of service, because I just couldn’t get to everyone. And that’s, that’s quite. It soon became quite clear that we, I, it wasn’t enough.” (an endometriosis nurse)*

Participants discussed being keen to improve their skills and knowledge (see theme A.4 for pressure for high knowledge levels). Participants reported being unable to attend conferences / training they felt would benefit the role.

> *“they won’t fund me to go to it this year [the BSG conference], which I think is a bit of a shame, because it’s not training that can be offered in house. […] if they want to improve women’s health, we need to go to these training courses” (an endometriosis nurse).*

#### Theme E: Skills and Boundaries

Some barriers and facilitators to the endometriosis nurse and PHWC roles were from the participants themselves. The skills and experiences people bring to the role can affect the service offered (such as whether certain tests or prescribing could be offered) and the motivation of the participant.

> *“I’m a non-medical prescriber, so I’ll be able to do prescribing within the community setting, and then also doing coils and then hormonal treatments as well” (an endometriosis nurse)*

##### E.1 Boundaries

Participants reported setting different boundaries, some in a less obvious way. Boundaries facilitated roles by protecting the service or the person’s own workload or wellbeing or knowing their own professional limits.

###### Boundaries for wellbeing

Participants discussed accepting and sharing that they needed a timeframe for email responses. Some participants reported avoiding or leaving online forums if their roles were discussed as that could have a negative impact, despite wanting to be there to understand patients perspectives. A participant described being able to limit her own workload by recognising that she was not running an emergency service, which allowed her to achieve a more sustainable balance.

> *“when I first started this job, I was probably working 54 hours plus a week. And at one point then I ended up being owed ridiculous amounts of hours to the point where I had to take a step back and think, ’[own name], this is ridiculous now. […] The one thing I emphasise is that I cannot be an emergency service. And that’s how I know I just have to switch off. I get back to them when I can, you know, and that’s all I can do. But as a nurse, I don’t feel right doing that. Does that make sense? It’s not what you want to be doing, but you have to look after your own mental sanity.” (an endometriosis nurse).*

###### Boundaries of professional limits

Participants explained knowing their professional limits.

> *“I know my boundaries, I wouldn’t just instantly start somebody on something [medication] that wasn’t, sort of, if I wasn’t sure” (an endometriosis nurse).*

Participants also discussed pushing back against service creep, such as a clinic that was meant to be used only for endometriosis patients becoming used by a wider variety of women.

###### Boundaries for service capacity

There was some discussion of limiting the service provided or publicised so that it did not become overwhelmed.

> *“We can’t take self referrals at the moment. We want to but I don’t think that we’ve got the capacity” (an endometriosis nurse).*

Participants reported having boundaries around their role from expectations of colleagues.

> *“you got to keep reminding him, I’m only here two days a week. And I’ve got, no one’s helping me with admin.” (a PHWC)*

#### Theme F: Context

##### F.1 Tertiary care

Not all health boards offer tertiary care and at least one does not have a district general hospital (DGH). The ease of access to tertiary care (specialist multi-disciplinary hospital care) and secondary care (district general hospital level care) differs and is a barrier to some participants.

> *“we’re having an awful time trying to refer our patients that need tertiary care into [tertiary care in another health board]. […] recently we’ve managed to get some funding approved […] to have, refer patients to [health board] […]But they’ve got their own long waits” (an endometriosis nurse).*

Participants working in a tertiary care centre report increased workloads as participants are expected to offer not just higher levels of care, but to patients from other health boards as well.

> *“We haven’t got the access and the time, the funded time to manage our patients. So we’ve now got more coming from [health board], and then we got ours as well.” (an endometriosis nurse)*

However not working in a tertiary centre can limit access to other disciplines.

*“[health board X] and [health board Y] are tertiary centres so they have to have established MDTs as part of their accreditation and we don’t have that, because we’re not an accredited centre” (an endometriois nurse)*

It can be seen that the different levels of care create more health board differences (see theme A.5). One participant explained that becoming a tertiary centre meant they would no longer need to reject referrals from other health boards.

> *“we can now open as a tertiary centre […] but obviously that’s going to take a lot of planning now and managing the finance. […It’s been] The downside of my job, and the worst part of my job, is telling somebody they can’t be seen in [health board] because they don’t live in [health board]” (a PHWC)*

##### F.3 Patient group

Participants empathised that many patients had been through a difficult journey, where healthcare professionals may not have listened to or even denied symptoms. Participants valued the time they could give patients and worked with understanding to help overcome these issues, as well as support ongoing symptoms whilst potentially on a very long waiting list.

> *“it’s mainly pain. And emotional as well, how they’re emotionally feeling, because it has a massive impact on their psychological state too. You know, I have had patients that have been suicidal because of the pain and we’ve had to get, like, the crisis team involved, because they really reached the end of their tether.” (an endometriosis nurse)*

Participants explained that expectations from services can be inaccurate and patients could be disappointed by the reality of what can be offered. Participants explained that they tried to be honest with patients about how their illness or symptoms could be managed.

> *“so they don’t want to keep seeing me because they just want to wait for their surgery. So that is really sad. [Interviewer: What do you feel like they were looking for?] A magic wand. I do say to patients when they first come into their appointment, you know, I haven’t got a magic wand. I’m not going to be able to solve everything, but I’m hoping that we can, if we can just help some of your symptoms to give you a better quality of life.” (an endometriosis nurse)*

A participant reported feeling that they had been subjected to abusive patient behaviours.

> *“I have had had a few abusive patients unfortunately and you get a few abusive texts and you’ve got no way of recording that. […] One of them was very serious threat, so it’s actually stopped me from doing, like, some charity work and stuff like that because of it, but, you know, you reflect and it was one person, which is horrible […] it’s just cause patients are angry, because they’ve been waiting so long. I, you know, I, sort of, understand it, but it’s not my fault as such, you know.” (an endometriosis nurse).*

##### F.4 COVID

During the COVID pandemic many healthcare services were stopped and staff re-assigned to emergency care and COVID-19 services. For services that stayed open or opened later in the pandemic some patients were too scared to attend [13]. This upheaval in the settings and beyond effected these newly developing women’s health services by disrupting the staff, but also engagement with the new service.

> *“before COVID we had a steering group and we had volunteer sector engagement. We had a really strong focus group really going. And it just all disbanded. And it’s really disappointing” (a PHWC)*

Participants explained a positive from the COVID epidemic in the acceptance and widespread use of telephone and online appointments for healthcare which some participants use frequently – especially in rural areas.

#### Theme G: Perceived Benefits of the roles

Participants discussed the benefits they felt came from the roles. As well as overt discussion, some themes were identified by researchers.

Benefits to patients, mainly from participants who were patient facing, stemmed from the time that these roles allow with patients as well as the high levels of expertise. The perceived benefits included:

- Reducing patient symptoms
- Improving patient quality of life
- Listening to patients often previously unheard journeys
- Reducing waiting lists
- Advocating, teaching and collaborating with others, including other healthcare professionals about their specialty and service.
- Improving their service

> *“We identified through the gynaecology department that patients that lived in [town] were travelling up to [hospital] to have their pessary changed. […] So we invited, we’ve got a practice nurse that runs a clinic on a Thursday with us and she sees all the pessary patients that live within the [town] cluster.” (a PHWC)*

There were also benefits to some of the participants themselves of being within these specialists roles.

> *“I’m very passionate about what I do, and I love my job. I’m very lucky that I’ve got a job that I love […] It’s the impact you have on patients. So it’s the difference that we can make from an admin role” (a PHWC)*

Some participants were able to share feedback they had received from patients.

> *“a lot of them say it’s just nice to have somebody to talk to that understands what they’re going through. […] it’s just nice to know that somebody understands what endometriosis is and that it’s not just painful periods. Because that’s what they’ve experienced before.” (an endometriosis nurse)*

> *“thank you for validating how I’m feeling and being that link between the consultant and the patient” (an endometriosis nurse)*

#### Theme H: Service development

Participants discussed previous service change and ideas they had for how their service could develop. There was mixed confidence in future ideas being realised. Participants discussed being constrained by resources and colleague support, so would need investment to expand. There is hope from some that the new Womens Health Plan [2] will support their services.

> *“the education within continence and pelvic health physios, they’re doing their own work, but that link with us all still needs to grow. And I think that will come with the Women’s Health Plan and the engagement from the All Wales level.” (a PHWC)*

##### H.1 Implemented or imminent changes

Roles have ‘*evolved’* since forming. Some services have expanded or improved, with many wanting to do more. One participant explained how they had improved the MDT and its attendance. In one health board, PHWCs discussed substantial changes led by a consultant (as a service advocate: theme B.1) of creating a centralised hub which had just become an Accredited Pelvic Floor Centre where services were relocated, encouraging more interprofessional working. In this centre, there were also new staff, a new pessary clinic, a ‘*tailor made*’ app to support patients and increased theatre time (through a business case). The PHWC is now also involved in a support group for patients.

In another health board the PHWC described expanding from one weekly consultant-led clinic to multiple nurse-led clinics in many sites, “*I just developed it and just made a, I don’t know, a success, I hope I have anyway. I think I have.*”

There were discussions from some PHWCs of working to improve referral pathways, including single point access and supporting patients accessing more conservative treatments while waiting for / instead of surgery, often following an audit.

> *“early on into the audit, that was the biggest thing we picked up on: Let’s get the patient to the right person, get the right care and then, surgery should always be the last port of call.” (a PHWC)*

##### H.2 Ideas for future service improvements

There was discussion from one endometriosis service proposing a one-stop clinic for women, though the participant reflected that service fragility (theme A.6) would be an issue:

> *“we’d like to set up, so, a one-stop nurse-led clinic, where the patient would come in, they would have clinical history taken, any investigations needed. […] So, the result would be given to that patient on that day. […] there’d be a plan of care at that appointment, which will reduce the footfall.” (an endometriosis nurse)*

There is a plan in one pelvic health service to add another surgeon and create a Wales-wide service.

##### H.3 Recommendations for the role

Improved connections to primary care

Endometriosis nurses in particular discussed reaching out to primary care and suggestions included:

- Support network for healthcare professionals
- Nurse-led endometriosis clinics within primary care clusters
- A named GP within each health board to support the roles

> *“All Wales Network[…] accessible to primary care, secondary care, and everybody can feed into it and seek advice from it and ask questions from it. It could be an open forum.” (an endometriosis nurse)*

###### Increasing awareness of and access to services

The public could be made more aware of pelvic health and endometriosis services / diagnoses:

- Signposting from other services
- Primary care promotions
- Education or promotions in schools
- Advertising and marketing around endometriosis and pelvic health
- “*more prompt and quick access to us.”* (an endometriosis nurse)

###### Improved colleague support and staffing

- Expanded MDT:*“I would just love to have physio and admins” (an endometriosis nurse) “psychological support.” (an endometriosis nurse)*
- A virtual women’s heath hub
- More admin support.
- Closer peer working: “*working autonomously, but working within a team” (endometriosis nurse)*
- Closer working between the PHWC and endometriosis nurse roles

> *“there’s this massive overlap from the two services, and we should probably be one big service, Pelvic Health Service, not endo services and urogynae” (a PHWC)*

- More PHWC and endometriosis staff which could then improve the referral pathway / support primary care / allow for cover / expand services. “*we need more endometriosis nurses*.” (an endometriosis nurse)

###### Increased knowledge sharing

- Videos for patients.
- Improving knowledge in primary care

> *“I think presenting and sharing knowledge is something that the next step for them [others in the role] is that they need to be going out and doing.”(an endometriosis nurse)*

###### Expanding the role remit

There was some discussion that the roles could support women beyond pelvic health and endometriosis: “*we want to expand, we wanna deliver as much as we can*.” (a PHWC)

> *“The plan, as far as I’m aware, is to expand to women’s health services. So it would go on to develop into sort of like menstrual cycle problems, heavy periods, and then on to menopause care as well. But we’re not there yet.” (an endometriosis nurse)*

### 3.3 Workshop Results

This workshop has not been thematically analysed, but summarized below into key messages.

#### Key difference to the interview data

Participants were asked to reflect on four themes from the interviews (see appendix three) and there was agreement (triangulation) that these themes remained important. However some discussion stood out as differing from the interview themes. There may have been a change since interview, or the workshop environment altered the discussion. In particular there was concern over increasingly difficult patient interactions, with some becoming aggressive. While participants understood the difficult time patients were going through and that they are ‘*crying out for help*’, the strain on participant’s own mental and physical health was too much and burnout was discussed. Some participants felt they could not walk away from aggressive / abusive patients as they were the only person carrying out the service.

#### Suggested ways forward

Participants discussed potential ways forward for the planned workshop themes, but also strayed onto other themes during the breakout session. These are summarised in figure five below.

**Figure 5:**
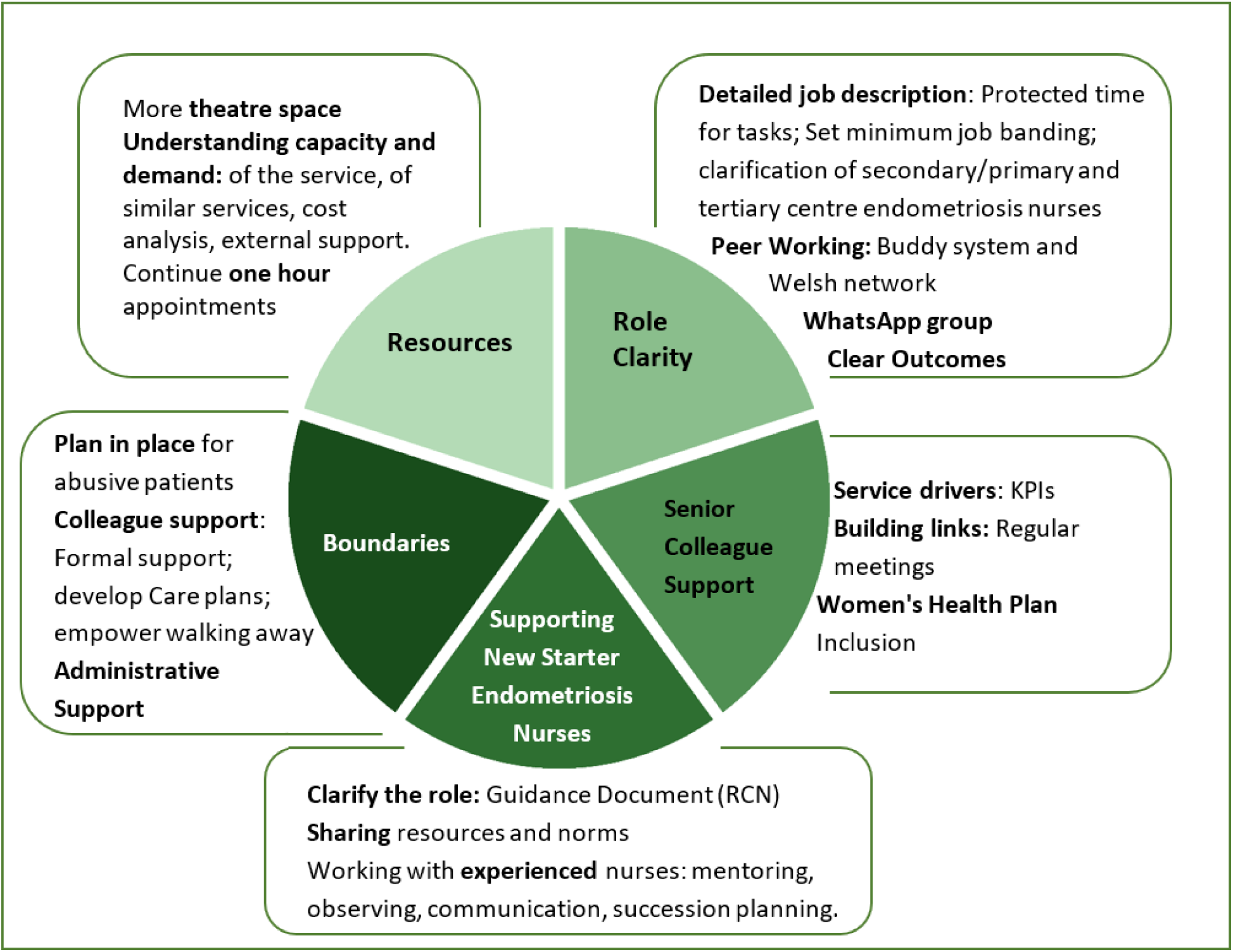
Suggested ways forward for themes discussed at the workshop

##### Resources

An understanding of the **capacity and demand** of these pelvic health and endometriosis services could support requests for funding. Also suggested was understanding similar services as a **benchmark** so that comparisons could be made of resource distribution.

##### Role clarity

Could be improved with detailed **job descriptions**. For PHWCs these may be different for different role types (eg clinical, administrative, managerial or governance). Minimum job banding, clarification of placement (secondary or tertiary care) and expectation of time for tasks would be useful. **Peer working** such as a buddy system, Welsh network and WhatsApp group could support role clarity. Clear **outcomes**, perhaps KPIs, could also increase role clarity.

##### Senior colleague support

Could be increased through service drivers such as **KPIs** as well as building **links**, for example regular meetings, with senior management. Participants felt they should be included in the **Women’s Health Plan** implementation or planning within their health board which could also support links with senior colleagues.

##### Supporting endometriosis nurse inductions

Participants reported supporting new starters but this could be more uniform. New starters should be signposted to the RCN **guidance** document for endometriosis nurses. There could be more (or more formalised) **sharing** of documentation and norms of the role. Working with **experienced** endometriosis nurses is important, such as being mentored and observing clinics. **Succession planning** is not currently resourced but is strongly reccomended by participants, it would also help overcome the fragility of the service.

##### Boundaries

More **administrative** support could help reduce workload and increase capacity. Discussion moved into how to support staff in these roles in the reported increasing problems of aggressive / abusive patients. Suggestions included, having a **plan** decided and in place ready for any issues.

**Colleague support** is important in coping with this as people in these roles are often the only one running a service and may feel they can not walk away from a difficult situation without support. One participant described that in case of an abusive interaction, she can leave the appointment and discuss the case at an MDT after which the patient can be sent a treatment plan. This means she is empowered to keep safe whilst knowing the team can still provide care.

## 4. Discussion

### 4.1 Summary of findings

Our in-depth qualitative study explored the lived experiences of people working as PHWCs and endometriosis nurses, and several factors that underpin the success of these roles. We also identified key barriers to carrying out the roles successfully and priority areas for change and improvement.

This service evaluation revealed that key factors in the success of these roles included support and working relationships from a supporting advocate to the MDT and senior leaders and primary care. Participants bring their own experiences to the role which can add value in different ways. Participants also create boundaries around their work or mental load to protect their wellbeing and / or service which, though not always stated overtly, was an important element of the role

Participants described barriers including their role description, especially its lack of clarity, with many differences across health boards. An outcome of this is that patients in health boards are likely receiving different services or care from the PHWC (in some health boards PHWC roles are not patient facing (e.g. administrative or governance roles). There can be difficulties getting started in the roles.

Resources and their availability effect the ability of participants and their service to carry out their core work, develop or allow staff to reach their potential. Reach and inclusivity of the roles is not well understood and views vary on this indicator. Interviewers reflected that participants found reach a more challenging question to answer and participants may have been more guarded in their response. Some participants felt that that self-referral routes for their services increased inclusivity but we are not able to understand from this service evaluation how well publicised, understood or easily found the self-referral routes are.

The workshop focused on the areas of role clarity, supporting new starters, colleague support, boundaries and resources. Key suggestions to support these roles moving forward (see figure five above and section 5 below) included: detailed job descriptions; improved peer working and resource sharing across Wales; clear outcomes; links with senior leadership; induction pack for new starters; succession planning; supporting boundaries, especially when dealing with abusive patients; more staff; administrative support; capacity and demand research of these and similar services; more research of the PHWC and endometriosis roles and to work more closely with consultants.

### 4.2 Our findings in relation to the Women’s Health Plan

The NHS Wales Womens Health Plan 2025-2035 [2] (WHP) aims to improve the health of women and girls across their lifetime, highlighting opportunities to reduce the gender gap. it includes short, medium and long term actions in eight priority areas, including endometriosis (priority two) and pelvic health and incontinence (priority five).

The WHP acknowledges the progress made in recruiting pelvic health and wellbeing co-ordinators and endometriosis nurses in each health board as well as setting up the Endometriosis Cymru website. It also acknowledges, as the participants in this study have raised “*that there is more to do to improve access and reduce variation across Wales.”* (Ministerial Foreword Page 6 [2]).

As experienced by participants in this study (resources theme D and patient group theme G.3) the WHP reflects there is a difficult journey for patients and “*evidence about women’s symptoms being undervalued, overlooked or dismissed*” (page 5, [2]) before looking at how that can be improved.

The WHP promotes evidence based healthcare and improved access to data for women in Wales. This aligns with the workshop suggestion that there needs to be further research into the cost benefit of these roles and of the capacity and demand of these services, in order to improve resource allocation.

Endometriosis and adenomyosis is priority area two within the WHP. The WHP recognises the importance of the introduction of the endometriosis nurses in Wales (page 53 [2]) and their impact. The action plan for priority area two (endometriosis and adenomyosis) is summarised in table five with reflections from the service evaluation.

Many of the WHP priority two actions align with endometriosis nurses’ suggestions and themes. However, participants seem to be raising more concern than documented in the WHP over:

- The fragility of the endometriosis service (only having one in post in most health boards, or per large geographical area).
- Lack of access to theatre for endometriosis patients creating long waiting lists.
- Rejection of referrals to allied healthcare professionals.
- A potential lack of reach and inclusivity of the service, though this needs data to ascertain.

The endometriosis nurse roles provide valuable time for patients to meet with passionate experts in their care, as stated at the start of the report:

> *“health boards should ensure there are appropriate levels of diagnostic, therapeutic and surgical capacity to enable women who require interventions for health needs specific to women and girls – including menstrual and fertility care, endometriosis and menopause – to receive care as close as possible to home without significant waits. (WHP page 6, [2])”*

Pelvic Health and Incontinence is Priority area 5 within the WHP and so relates to the PHWC role. The WHP defines Pelvic floor dysfunction as: “*an umbrella term encompassing a wide range of conditions in which the pelvic floor muscles around the bladder, anal canal, and vagina do not work properly. The three most common and definable symptoms of pelvic floor dysfunction are urinary incontinence, pelvic organ prolapse and faecal incontinence. However, others include emptying disorders of the bladder and bowel, sexual dysfunction and chronic pelvic pain*.” ([2] page 64) and effecting 60% of UK women.

**Table 5:**
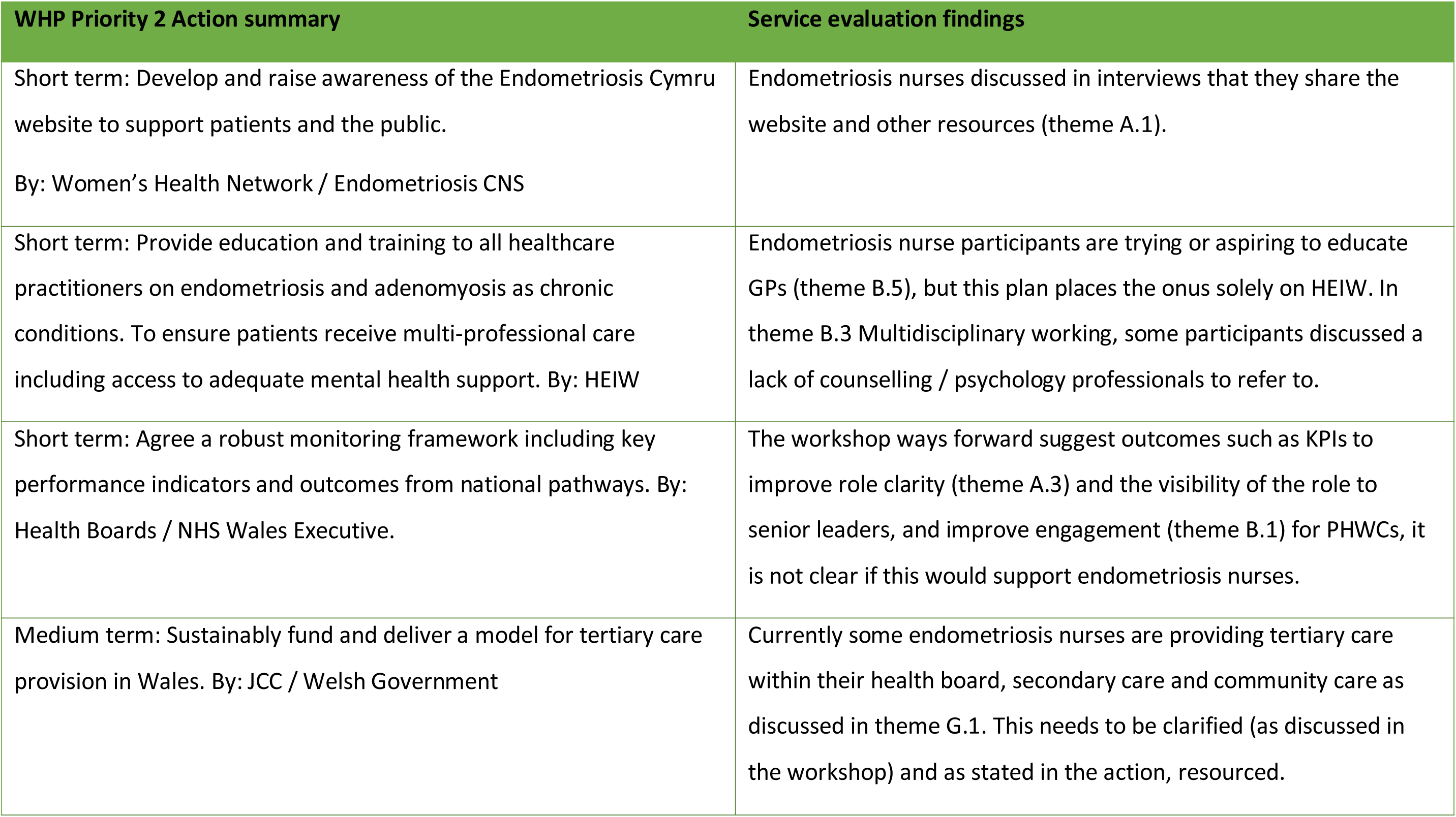
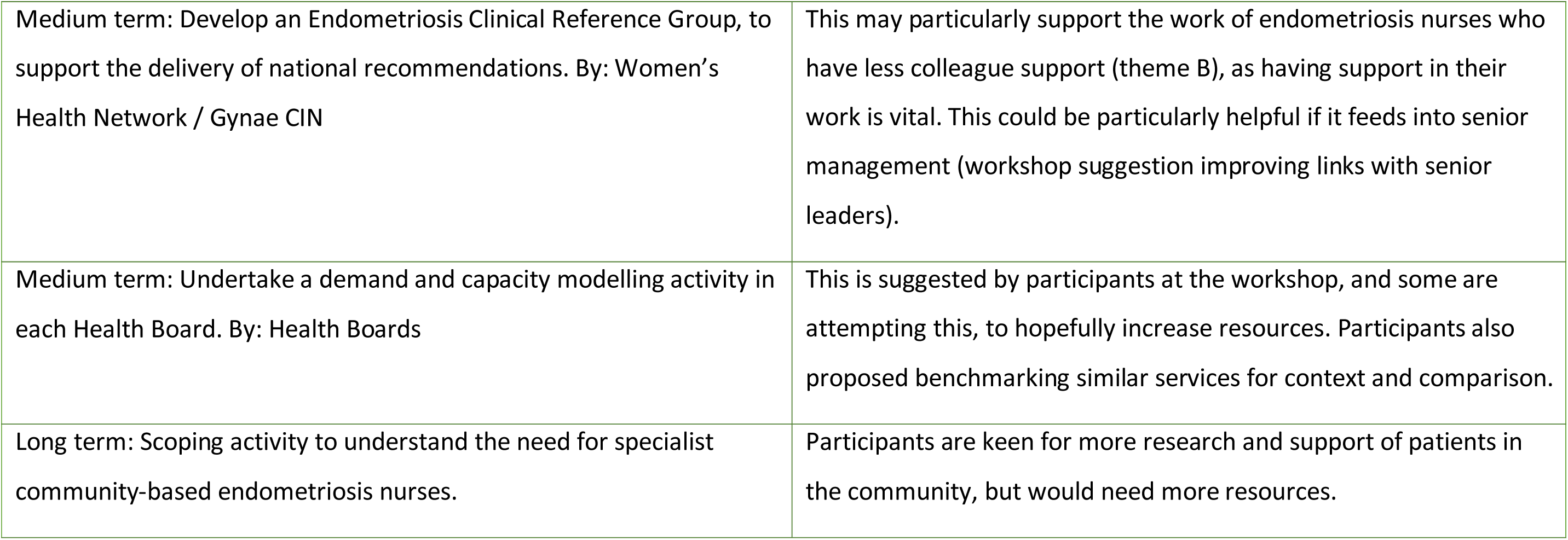
The WHP endometriosis and adenomyosis action plan with service evaluation findings –summarized from the WHP [2] page 55.

| WHP Priority 2 Action summary | Service evaluation findings |
| --- | --- |
| Short term: Develop and raise awareness of the Endometriosis Cymru website to support patients and the public.<br>By: Women’s Health Network / Endometriosis CNS | Endometriosis nurses discussed in interviews that they share the website and other resources (theme A.1). |
| Short term: Provide education and training to all healthcare practitioners on endometriosis and adenomyosis as chronic conditions. To ensure patients receive multi-professional care including access to adequate mental health support. By: HEIW | Endometriosis nurse participants are trying or aspiring to educate GPs (theme B.5), but this plan places the onus solely on HEIW. In theme B.3 Multidisciplinary working, some participants discussed a lack of counselling / psychology professionals to refer to. |
| Short term: Agree a robust monitoring framework including key performance indicators and outcomes from national pathways. By: Health Boards / NHS Wales Executive. | The workshop ways forward suggest outcomes such as KPIs to improve role clarity (theme A.3) and the visibility of the role to senior leaders, and improve engagement (theme B.1) for PHWCs, it is not clear if this would support endometriosis nurses. |
| Medium term: Sustainably fund and deliver a model for tertiary care provision in Wales. By: JCC / Welsh Government | Currently some endometriosis nurses are providing tertiary care within their health board, secondary care and community care as discussed in theme G.1. This needs to be clarified (as discussed in the workshop) and as stated in the action, resourced. |
| Medium term: Develop an Endometriosis Clinical Reference Group, to support the delivery of national recommendations. By: Women's Health Network / Gynae CIN | This may particularly support the work of endometriosis nurses who have less colleague support (theme B), as having support in their work is vital. This could be particularly helpful if it feeds into senior management (workshop suggestion improving links with senior leaders). |
| Medium term: Undertake a demand and capacity modelling activity in each Health Board. By: Health Boards | This is suggested by participants at the workshop, and some are attempting this, to hopefully increase resources. Participants also proposed benchmarking similar services for context and comparison. |
| Long term: Scoping activity to understand the need for specialist community-based endometriosis nurses. | Participants are keen for more research and support of patients in the community, but would need more resources. |

### 4.3 Strengths and limitations

#### Strengths

A key strength of this service evaluation was the successful recruitment of all eligible participants for interviews. This gave the maximal insights into the roles being evaluated. Additionally, there was strong engagement at the workshop, with high attendance and active, thoughtful contributions from participants. These high levels of engagement were largely due to the willingness of individuals in these roles to share their experiences and contribute to improving the service.

#### Limitations

The data presented here is not generalisable beyond this specific context. They reflect the perspectives of individuals in particular roles within Wales at a specific point in time, which is consistent with the nature of a service evaluation. While some of the lessons learned may be transferable to other roles or settings, they should be interpreted with caution. This service evaluation also only shows one perspective: of those in the role. This is an important perspective but does not give the entire picture of the PHWC and endometriosis nurse services, where patient perspectives really supplement the narrative as well as those who refer to, take referrals from, line manage, and oversee the roles.

### 4.4 Conclusions

Participants in these highly specialist roles demonstrated strong commitment and passion for their work, despite facing a number of barriers to effective delivery. They also identified key facilitators that supported their roles—most notably, the support and collaboration of colleagues.

## 5. Implications for practice and policy

As well as suggestions from the workshop (section 3.3), there are interview and researcher team suggestions of how the PHWC and endometriosis nurse roles could be supported, and outlined in table 6:

**Table 6:**
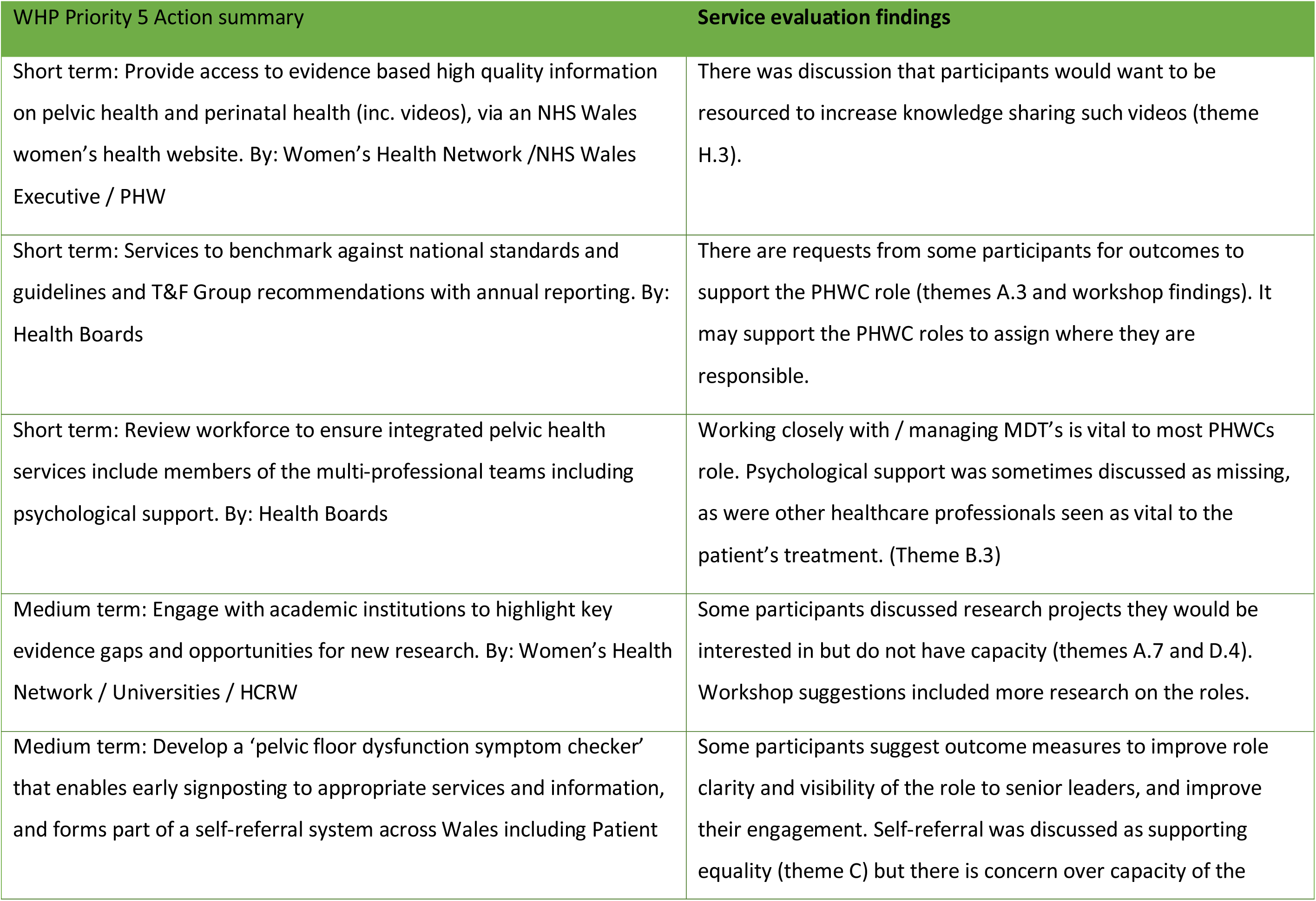
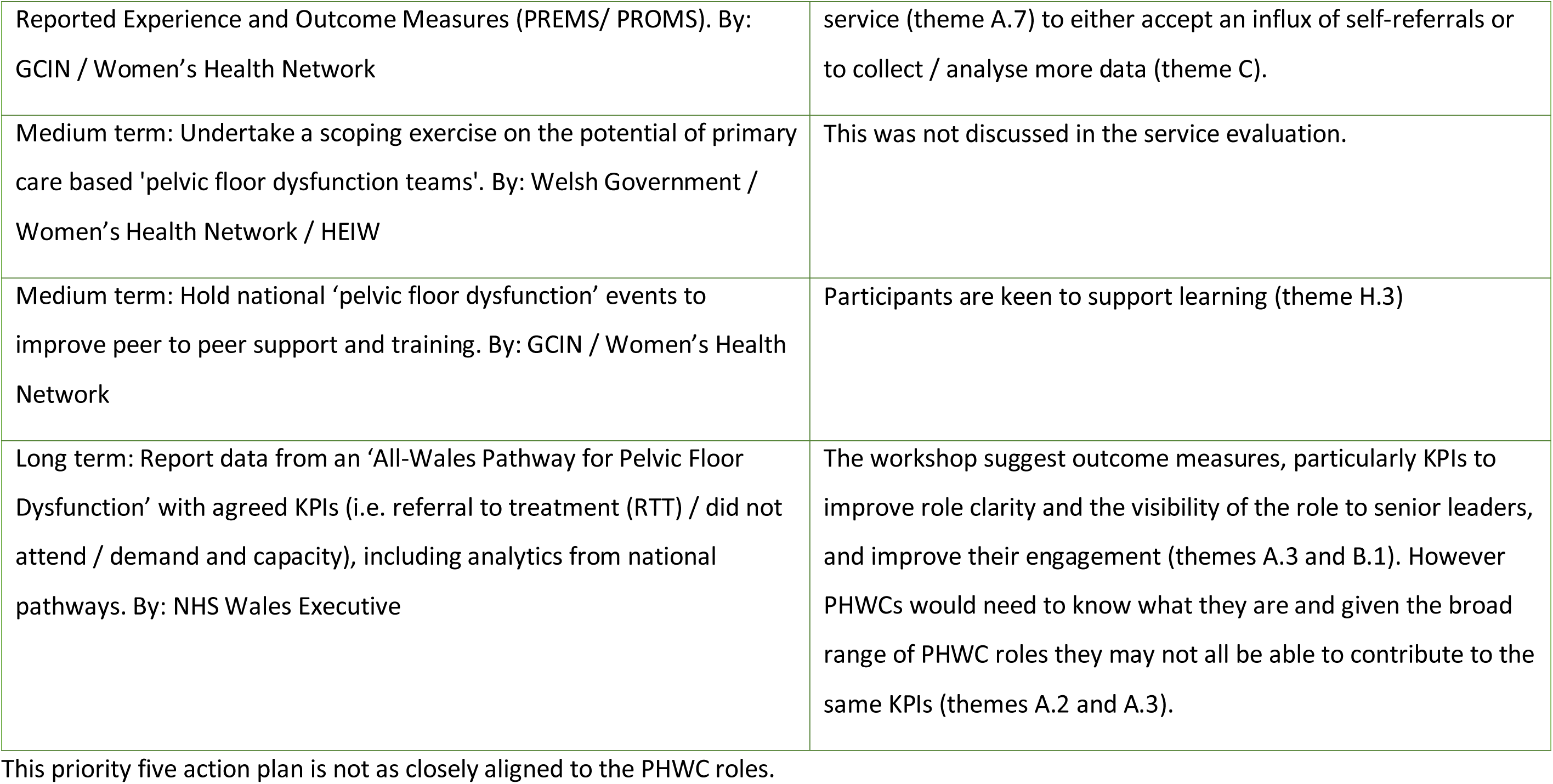
The WHP pelvic health and incontinence action plan with service evaluation findings –summarized from the WHP [2] page 66.

**Table 7:** Suggested potential improvements to the PHWC and endometriosis nurse roles.

| Suggested Improvement to the Role(s) | Reason for suggestion | Source |
| --- | --- | --- |
| More resources: Theatre time for endometriosis and pelvic health patients; more staff; administrative support. | Reduction in waiting lists from increased theatre time would support patients, the service and staff experience. Increased staffing and administration support would increase capacity and cover of staff and the service for their primary responsibilities. It could improve service reach. | Workshop, interviews |
| Detailed job descriptions including: expected time for different tasks, minimum banding, distinguish different sub-roles, e.g. tertiary endometriosis nurses and different PHWC roles. | Improve role clarity and support boundaries. | Workshop |
| Improved peer working across Wales. Could include: (1) WhatsApp group, quickly adding new starters. (2) Regular online meetings with a specific learning focus. With either a single or rotating lead to organise each meeting. (3) Co-operative online space for resources including funding ideas. (4) Learning collaborative. | Improve role clarity, support for new starters, improved support, opportunities for learning and sharing funding ideas for more potential resources. | Workshop, Interviews |
| Introduction of outcome measurements such as KPIs. With flexibility for the variety of PHWCs who may need differing outcomes. This is particularly important for the PHWC role. | Encourage senior health board engagement.<br>Improve role clarity. | Workshop,<br>interviews |
| Capacity and demand evaluations: Also benchmarking similar services. | To support bids or business cases for more resources. | Workshop |
| Plan in place in case of an abusive patient as participants report an increase in cases. Some did not have a plan and some single providers of a service felt unable to respond. | Staff in these roles need to be able to identify when a patient has become abusive and know what to do, with colleague support, in advance, for their safety. | Workshop |
| New starter endometriosis nurse induction pack. Including links to shared resources, RCN guidance document, Fair Treatment For Women in Wales (FTWW) endometriosis nurse patient-led toolkit, ideas for how to get started (e.g. observations of clinics etc.). Also could be considered: (1) a tool to recognise readiness to start independent clinics (2) create a simple training video (3) assigning a mentor. | To support new starter nurses to begin their preparation more quickly.<br>Encourage peer working from the start of the role | Developed<br>from the<br>workshop |
| Acceptance of referrals by other healthcare professionals | Improved MDT working |  |
| Inclusion in the planning, implementation and evaluation of the Women's Health Plan (WHP). | Encourage senior colleague support and role visibility<br>Support service change through the WHP | Workshop |
| Improved primary care relations such as through a named GP contact, perhaps a designated GP Cluster lead for each health board. | Improved colleague support | Interviews |
| Guidance for the endometriosis nurse voicemail / inbox to support staff to make boundaries. | Improve staff understanding of boundaries around this task to support their own capacity | Researcher, interviews |
| Training for staff in self-management support: when a healthcare professional supports a patient with feeling more competent in managing their life with illness[14]. | This may support staff confidence, empower patients. It may improve capacity as empowered patients may be less reliant on their healthcare professional. | Researcher |
| Promotion of the pelvic health and endometriosis nurse services once there is capacity. Such as education in schools, signposting from other services and advertising. | This would improve reach of the service though care must be taken that any inequalities are not exacerbated. | Interviews |
| Succession planning | Having a plan for training the next person for the role could ease pressure on those in the role. A person in training may be able to partly cover for the lead staff member during periods of leave. | Workshop |
| Support with funding | Support with funding sources and applications may support their work, though capacity will be an issue. | Interviews. |
| Addition of necessary staff to the MDT to support the work of the pelvic / endometriosis service, such as psychological support, pain team where needed. | This would improve the support for patients and staff. | Interviews |
| Improved senior colleague support such as closer working with consultants and senior management | This could improve role visibility, impact of the role and clarify messaging to patients. | Interviews, workshop. |

## Data Availability

All data produced in the present study are available upon reasonable request to the authors

## Abbreviations

PHWC: Pelvic Health and Wellbeing Coordinator
WHIG: Women’s Health Implementation Group
MDT: Multi-disciplinary team
KPI: Key Performance Indicator
FTWW: Fair Treatment for the Women of Wales
HCP: Healthcare Professional
BSGE: The British Society for Gynaecological Endoscopy
WHP: Women’s Health Plan
DGH: District General Hospital
RCN: Royal College of Nursing

## 7. Acknowledkgements

The authors would like to thank the endometriosis nurse and pelvic health and wellbeing coordinators who participated in the interviews and workshop for this service evaluation who gave their time and experiences so graciously.

## 8. APPENDIX

## Appendix 1 Interview schedule

**The guide has been underpinned by the Implementation Outcomes Framework and the MRC Process Evaluation Framework.**

It is semi-structured so will be used differently for each participant. It is also iterative and will be reviewed throughout the study.

### Introduction

1. **Introduce yourself**, thank participant, explain where you are calling from. Confirm time participant needs to finish by.
2. **Set the focus of the interview/ Study description:** I am working in the Health and Care Research Wales Evidence Centre. We are currently speaking to Pelvic Health Co-ordinators and Endometriosis Nurses across Wales to see how these relatively new services are running, what is going well and what barriers are being faced. We hope this may be an opportunity to enhance the services by continuing to improve and learn from each other as well as support other women’s health services in Wales. We are keen to hear directly from you, the nurse (or co-ordinator), as we really value your experience and knowledge from doing the work. I would like to reassure you, there is no need to worry as there are no right or wrong answers and the outcome will not be used against you in anyway. This is a safe space.
3. **Check understanding** and answer any questions. Does that make sense? Do have any questions about the study or about why I’m speaking to you today?
4. **Consent** discussion I can see you have completed the electronic consent form (X days ago). Are you happy you understood the form? [*answer any queries*] I just want to remind you that we can stop whenever you like, if you want a break or if you want to completely stop the interview that is fine. We will be recording but I will let you know when that starts. Just because you have signed the consent form does not mean you have to finish if you do not want to, so please do let me know if you change your mind.

**Signposting in case the participant becomes distressed during interview**

Advice to contact their GP in the first instance

**Canopi** offers a free and confidential mental health support service for social care and NHS staff in Wales aged 18 years and over. Offer various levels of mental health support including: Self-help; Support from Wellbeing Allies; Guided self-help; Virtual and face-to-face therapies with accredited specialists. **0800 058 2738.** https://canopi.nhs.wales/contact-us/

**Healthcare workers foundation:** charity founded by healthcare workers, for healthcare workers, to address crucial welfare and wellbeing needs. Provide help in the form of financial support, counselling and bereaved family support. **0203 576 0374** https://healthcareworkersfoundation.org/

I will now be starting the recording. **START RECORDING**

#### 1. About your role

1. **Tell me about your role.**

a. Can you explain your day-today activities in work?
b. Can you tell me about the aims of your role?
c. How do you feel about the role?
d. Do you have another NHS role? How does this work?
2. **How did you prepare for this role?**

a. Prompt: did you do any specific training /mentoring/previous work experience?
3. Can you tell me about the **support** you are given in your role?

a. Prompts: Patient, clinician, team, other staff, administrative, organisation, structural, policy, attitude of people around you, training, supervisor.
4. We are trying to understand how the system works, what happens after you have seen a patient?

a. Talk me through the onwards referral process if they are needed.
b. Do you need to refer back to a GP or directly refer on to appropriate teams?

#### 2. Working with others

1. **Do you work alongside other practitioners as part of this service?**

a. How do you work together to achieve the aims of your service?
2. How would you describe the attitudes of your colleagues (and the organisation) towards your role?
3. What are your colleagues (and the organisation’s) expectations of your role?

a. How do you feel about that?

#### 3. Patient Experience

1. **Do you feel that the right patients are being reached?**

a. How are patients identified or referred?
b. How could this be improved? Has it been already?
2. **Do you think your role has had an impact on your patients?**

a. What makes you think this?
b. Do you feel you are seeing people from different backgrounds?
c. Do you think they are benefitting equally?
3. Do you feel your role meets the needs of your patients?

a. How could that be improved?
b. What (if anything) have you changed to support them?
4. Have you had any patient feedback relating to the service? Both Positive & neg
5. What else do you think patients want from the service?

1. As far as you are able to say, are patients able to contact the service in the way they want to, at the time they want to?
2. Do you feel there have been any unintended consequences of your role?

#### 3. Changes to the role

1. Has your role evolved since startingHow? Why?

a. Explore why: improvements vs barriers
b. If no: Do you think your role needs to or should change?
2. **Have you found anything you think could be improved for your role?**
3. Has anything **hindered** (or acted as barriers) your being able to carry out your role as intended? (patient, clinician, team, organisation, structural, policy)
4. If you have experienced any support/barriers, have there been any differences across different people or **settings** you are based in?

a. What do you think might have contributed to this? E.g. attitudes, roles, institutional factors?

#### 5. Recommendations

1. **What would be your key recommendations for how your role could improve?**
2. What has worked well in your role? Why?
3. What have been the most significant challenges?

a. How could/have these be overcome?
4. Since your role has started, what do you think has been the most significant change?

END: Thank you for your time today

I am going to turn the recorder off, please stay for a moment.

##### RECORDER OFF

Debrief participant unrecorded

## Appendix 2 Interview consent form

### The views of Endometriosis nurses and Pelvic Health & Wellbeing Co-ordinators

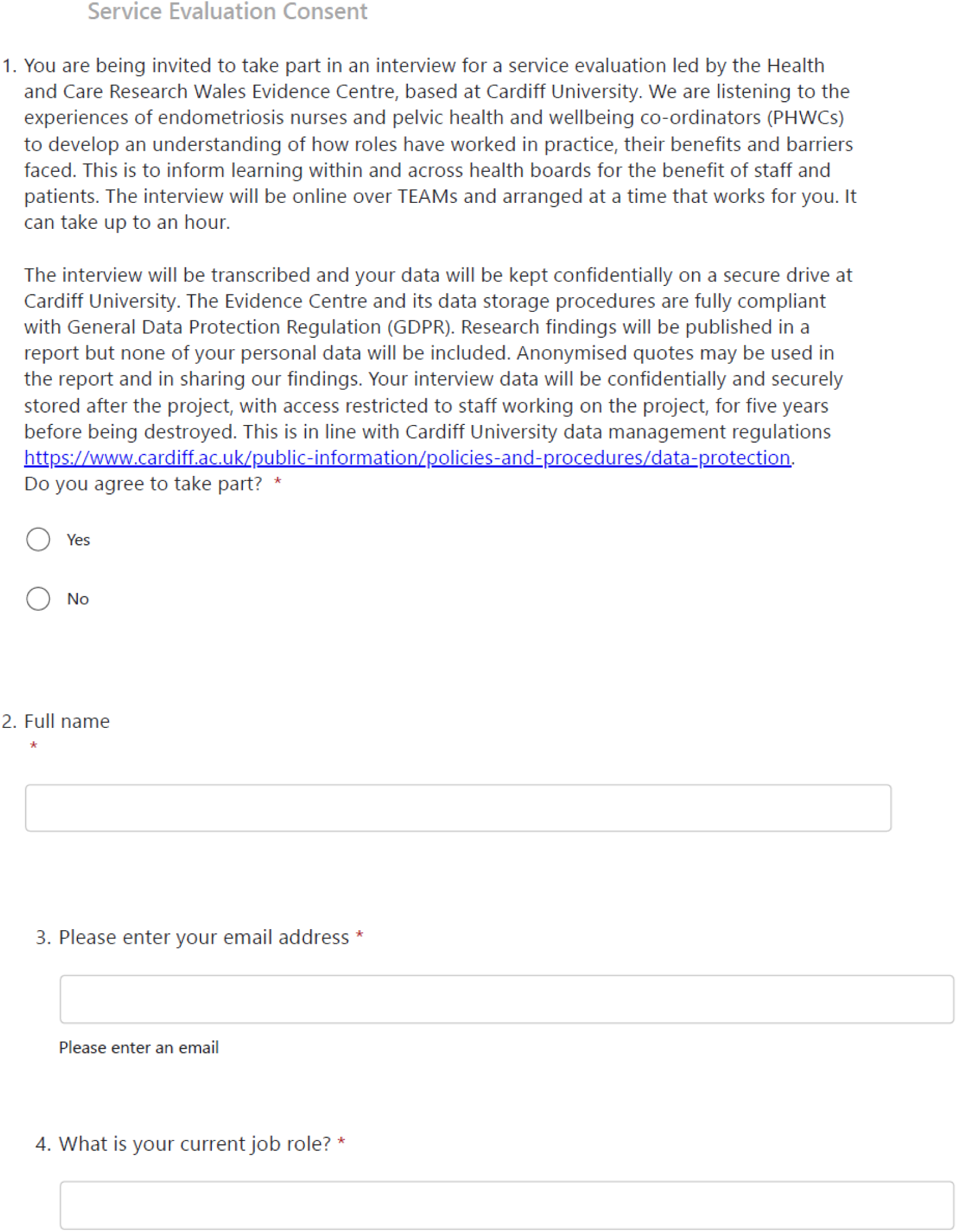
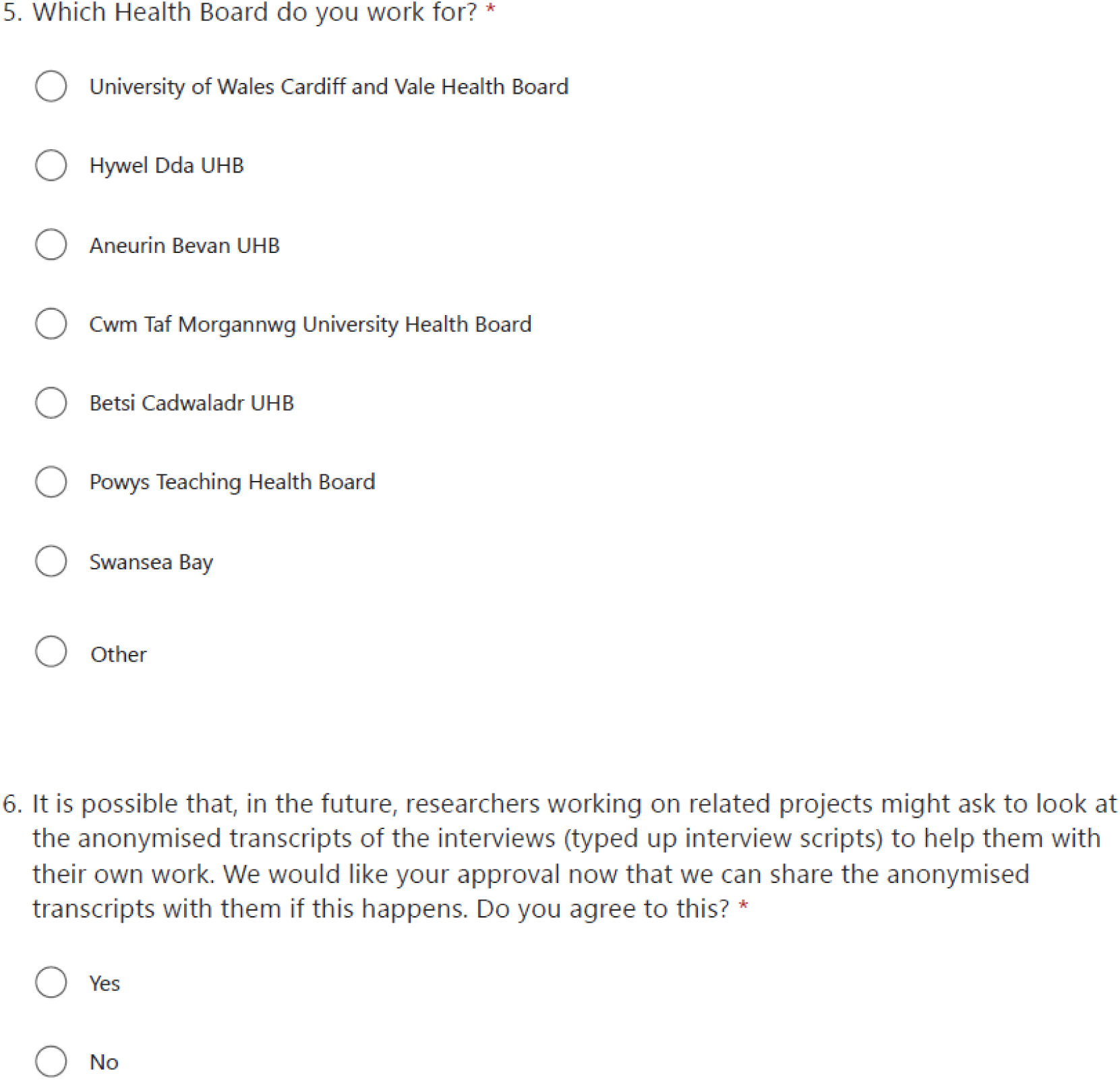

## Appendix 3 Workshop Breakout rooms Facilitator Guide

### Pelvic Health Wellbeing Coordinator Room topics: Facilitator: DW and EC

1) **Role clarity** and service variation: the co-ordinator role is really varied

a) Are sub-roles helpful to clarify the role further? Such as those considered in this evaluation (eg, clinical, administrative, managerial and governance).
b) What else may be helpful? For those in role and working with colleagues to define the role so that people understand it.
c) What are the must-haves for the role? What is the warranted variation?
2) **Resources and finance** often restrict the work, particularly administrative support:

a) There are other potential sources of funding, eg health board, what support is needed to apply for and potentially capture these?
b) How do you look for finance support?
**3) Colleague Support:**

a) How can we improve relationships with or expectations from colleagues?

i) This includes other healthcare professionals within the setting, or others in the same role across Wales, line managers or seniors within trusts.
b) How can those who may not have a person working with them that can help drive change be better supported?
c) What supports MDT working?
4) **Boundaries** at work:

a) How can useful boundaries at work be developed?
b) What do pelvic health wellbeing coordinators need to be able to create boundaries that enable their work while being flexible for the needs of the health board?

### Endometriosis Nurse Room topics: Facilitator: AC and NR

1. How can we support **new starter** endometriosis nurses?

a. Would a guidance document help? How could it be generated and what could it include?
b. How could a new starter know when they are ready to go, for example to run clinics?
2. **Colleague Support:**

a. How can we improve relationships with, or expectations from colleagues?
b. How can we support those who may not have a person working with them that can help drive change?
c. Could there be a potential for international or national connections such as a learning collective which brings together people interested in endometriosis, to share learning, links, resources etc.

1. **Boundaries** at work:

a. How can we support each other in creating useful boundaries at work?
b. There was a lot of discussion of the open access email inbox or call lines for endometriosis nurses as creating a lot of work that could be overwhelming, creating boundaries around this may be a way of supporting a sustainable way forward. What kind of boundaries could support this ongoing patient access?
2. **Resources and finance** often restrict the work, particularly administrative support:

a. There are other potential sources of funding, eg health board, what support is needed to apply for and potentially capture these?
b. How do you look for finance support?

## Appendix 4 Workshop invite

Dear [name]

We are pleased to invite you to an **online** workshop for the Womens Health: Endometriosis Nurses and Pelvic Health and Wellbeing Coordinators Service Evaluation on **Tuesday 24^th^ June at 12:00 - 13:30pm.**

We have planned it over lunch time in the hopes that it will make it easier for you to attend.

During the workshop, we will summarise findings to date and get your insight into the implications for policy and practice going forward.

You will shortly receive a calendar invite - **please RSVP to the calendar invite** so we know who to expect and so you receive the placeholder in your calendar.

We do hope you can join us. Further information about the workshop is below, but please do let us know if you have any questions.

Best wishes

Elly Clarke

**More detailed information about the workshop:**

### What will happen at the workshop?

The Evidence Centre team will present the findings from interviews with pelvic health and wellbeing coordinators and endometriosis nurses, highlighting the key themes. We will then break into groups to discuss potential solutions and ways forward on selected implications for policy and practice.

### Do I have to come?

You do not have to come. We know from speaking to participants at interview that a lot of you would like to come. If it works with your schedule and we would like to hear from you, but there is no pressure at all to attend.

### Will it be anonymous?

You will be discussing your view with other participants during the workshop. However, your contributions will remain anonymous in our reporting. We would like people who attend to have their cameras on, as that helps us to converse and see when people are talking. At the start of the session we will set the expectation that we will not discuss the content of the workshop outside of the workshop. When we put any results of the workshop into the report we will not use names or identifying information.

### How will the workshop be used for the project?

We plan to use the workshop to refine our policy and practice implications, which will be based on the themes of the interviews with endometriosis nurses and pelvic health and wellbeing coordinators. We plan to record the workshop, so that we do not have to take full notes and can listen to you. This will help us in case we need to go back to the recording to check anything. We may use anonymous quotes (that do not identify the speakers) from the workshop to describe the policy implications and potential ways forward.

### Who else will be there?

We are currently finalising who would best be invited to the workshop, but we will are inviting pelvic health and wellbeing coordinators, endometriosis nurses, public partners (including Fair Treatment for Women in Wales) and representatives from Welsh government who are stakeholders in the work.

